# Endemic means change as SARS-CoV-2 evolves

**DOI:** 10.1101/2023.09.28.23296264

**Authors:** Sarah P. Otto, Ailene MacPherson, Caroline Colijn

## Abstract

COVID-19 has become endemic, with dynamics that reflect the waning of immunity and re-exposure, by contrast to the epidemic phase driven by exposure in immunologically naïve populations. Endemic does not, however, mean constant. Further evolution of SARS-CoV-2, as well as changes in behaviour and public health policy, continue to play a major role in the endemic load of disease and mortality. In this paper, we analyse evolutionary models to explore the impact that newly arising variants can have on the short-term and longer-term endemic load, characterizing how these impacts depend on the transmission and immunological properties of variants. We describe how evolutionary changes in the virus will increase the endemic load most for persistently immune-escape variants, by an intermediate amount for more transmissible variants, and least for transiently immune-escape variants. Balancing the tendency for evolution to favour variants that increase the endemic load, we explore the impact of vaccination strategies and non-pharmaceutical interventions (NPIs) that can counter these increases in the impact of disease. We end with some open questions about the future of COVID-19 as an endemic disease.

## Introduction

Early in the global pandemic, COVID-19 levels rose and fell steeply, displaying rapid exponential growth and leading to widespread lockdowns and other public health measures to slow transmission (Ogden et al. 2022; Talic et al. 2021). The vaccination campaigns of 2021, followed by the nearly-uncontrolled Omicron waves in early 2022 (Figure 1, BA.1 and BA.2 peaks), have now led to almost 100% immunological exposure in many countries. In Canada, for example, 100% of blood donors had developed antibodies to the spike protein from previous exposure to the virus by June 2023, with 80% also showing antibodies to nucleocapsid, indicating prior infection (Canadian Blood Services 2023). The number of immunologically naïve individuals that fed COVID-19 dynamics throughout the pandemic has now greatly decreased, but in its place is a continual flow of newly susceptible individuals as humoral immunity wanes. For the past year, COVID-19 levels have ebbed and flowed in response to this waning immunity, new variants, and to changing public health measures. These peaks and troughs are more subdued wavelets, compared to earlier Omicron peaks (Figure 1).

**FIGURE 1:**
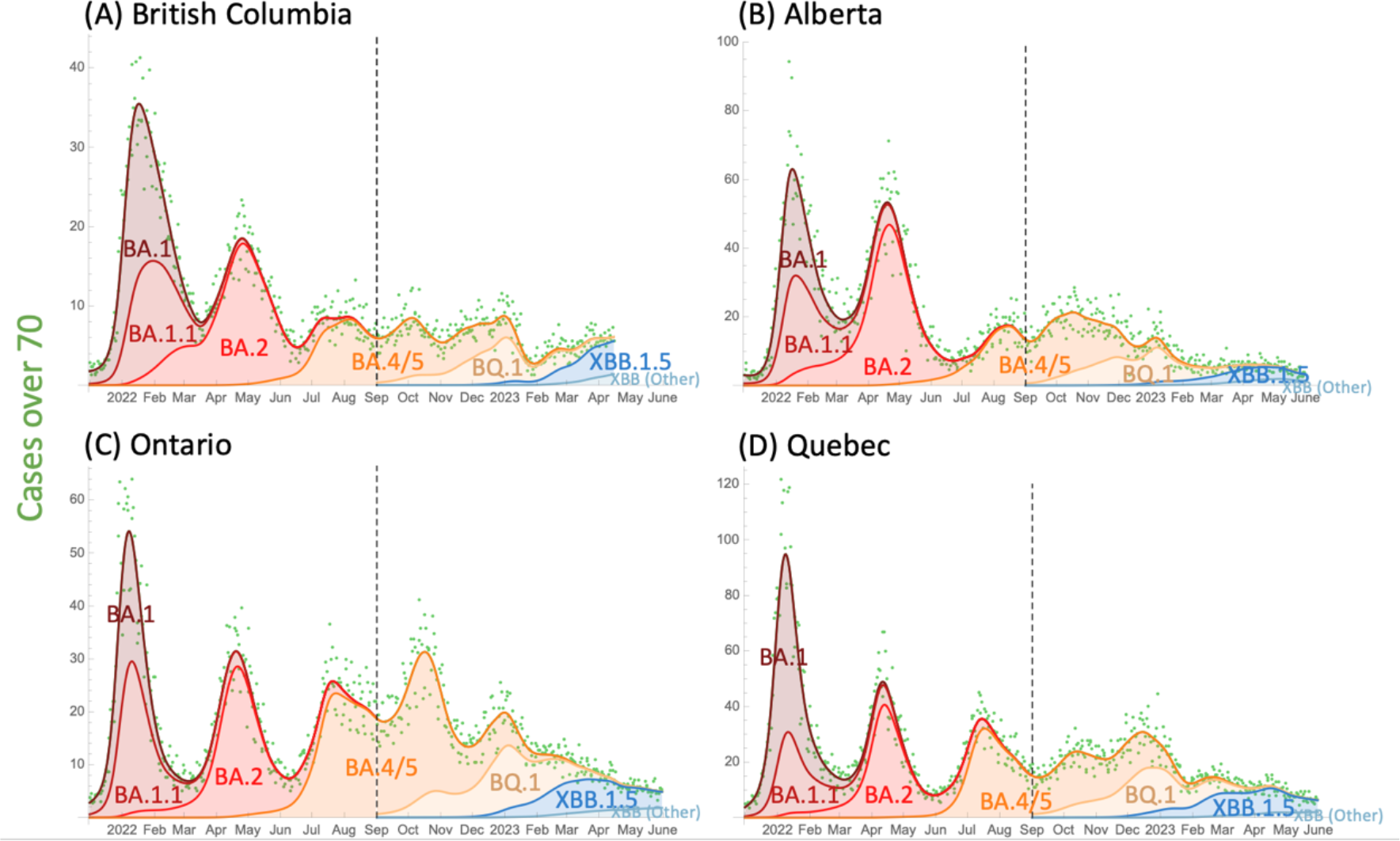
COVID-19 trends across four provinces in Canada. Major waves in early 2022 were driven by the rise and spread of Omicron, whose immune-evasive properties allowed widespread infection at a time when public health measures were largely relaxed (peak in January 2022: BA.1, April: BA.2, July: BA.4 & BA.5). A year later, Omicron variants have continued to spread rapidly (peak in December 2022: BQ.1; April 2023: XBB.1.5), but they no longer cause major waves in cases. PCR-confirmed cases per 100,000 individuals aged 70+ (green dots) are used to illustrate case trends, as testing practices changed dramatically over this time period but this age group remained eligible for testing. To guide the eye, a cubic spline fit (lambda =3) was applied (top curves in each panel), and the frequency changes of each variant under this curve were fitted by maximum likelihood using duotang (CoVaRR-Net’s CAMEO 2023). Genomic sequence data from each province were obtained from the Canadian VirusSeq Portal (VirusSeq 2023) and fit by maximum likelihood to a model of selection in two periods: first 9 months using BA.1 as a reference (left of dashed line); second 9 months using BA.4/5 as a reference (right of dashed line), grouping all clades within a family together except when a sub-clade is also mentioned (e.g., BQ.1 separated from BA.5). See supplementary *Mathematica* file for scripts and duotang (CoVaRR-Net’s CAMEO 2023) for methodological details and finer resolution of lineages and time periods.

COVID-19 is now considered an endemic disease, being both widespread and persistent, adding to the respiratory infectious diseases with which we must routinely contend. Its now-endemic nature reflects a balance between waning immunity and on-going transmission, leading to a turnover of cases across the globe. Endemic does not mean “constant”, as new variants and behavioral shifts drive change. Endemic also does not mean *“*rare”, as waning and transmission rates have remained high (e.g., Figure 1). Here we explore mathematical models to improve understanding of how the ongoing evolution of SARS-CoV-2, as well as our behavioural responses, will shape endemic COVID-19 and similar diseases.

When most individuals in a population are susceptible (epidemic phase), any variant or behavioural measure that affects the transmission rate will have a direct effect on the number of new infections over the short term, as exposures determine the spread of disease. When a disease first appears, the reproductive number describing the number of new infections per infection, *R*_0_, is given by the transmission rate divided by the clearance rate of the infection in the classic SIR epidemiological model (Keeling and Rohani 2011). Thus, variants that increase transmission or behavioural changes that reduce transmission directly reduce new infections and the rate of exponential growth, but these new infections have little immediate effect on the large pool of susceptible individuals. Models of this epidemic phase (e.g., (Day et al. 2020)) also typically ignore waning of immunity or the possibility that variants may evade any such immunity earlier.

By contrast, when a disease is endemic and most individuals have some degree of immunity, waning must be explicitly considered and modeled in a manner that allows variants to infect earlier (as in the SIR_n_ model, with multiple recovered classes, that we consider here). Furthermore, any change in the transmission rate has a more complex effect on the number of infections in the near future, because higher transmissibility depletes the number of susceptible individuals whose immunity has waned (by “refreshing” that immunity through exposure), while lowering transmission allows susceptible individuals to accumulate.

Indeed, this kind of reasoning about endemic disease has been used as an argument against non-pharmaceutical measures like masking:

> Masks “can delay transmission, they can reduce transmission, but they’re not actually effective measures at a population level,” because “exposure is essentially universal now to COVID-19.”
>
> Public Health Officer, November 2, 2022 in <u>Today in BC podcast</u>

This argument assumes that transmission is so common that reducing risks of exposure no longer matters, as another exposure will occur soon thereafter. This is a strong claim, with major implications for both individual and public health decisions. It is essentially a claim that endemic levels of a disease such as COVID-19 are not under our control.

Mathematical models can help evaluate such claims and determine whether and to what extent our actions affect the endemic load of disease and mortality. Models can also predict how this load would change in the face of new variants and how this depends on the properties of those variants. Here, we tailor standard epidemiological models to the current phase of COVID-19 to better understand the risks posed by new variants and our ability to control endemic diseases.

### Model background

We use a classic compartment model, SIR_n_, as illustrated in Figure 2, measuring the fraction of the population that is in the susceptible class (*S*), the infectious class (*I*), or in one of several recovered classes (*R*_*j*_). These sequential recovered classes allow for different stages of waning immunity (*i*, ranging from 1 to *n*) and capture the dynamics of neutralizing antibodies that help protect against infection (Andrews et al. 2022; Khoury et al. 2021). When measured on a log scale, neutralizing antibodies rise to a high level soon after infection or vaccination and then decline linearly over time since vaccination and/or infection (e.g., (E. H. Lau et al. 2021; Evans et al. 2022; C. S. Lau et al. 2022; Jacobsen et al. 2023). We thus consider the *R*_*j*_ classes to fall along different stages of this linear decline (*R*_1_ being highest and *R*_*n*_ lowest), with antibody levels falling over time (modeled as movement of individuals from *R*_*j*_ to *R*_*j*+1_) until levels are so low that infection is no longer prevented (*R*_n_ waning to *S*).

**FIGURE 2:**
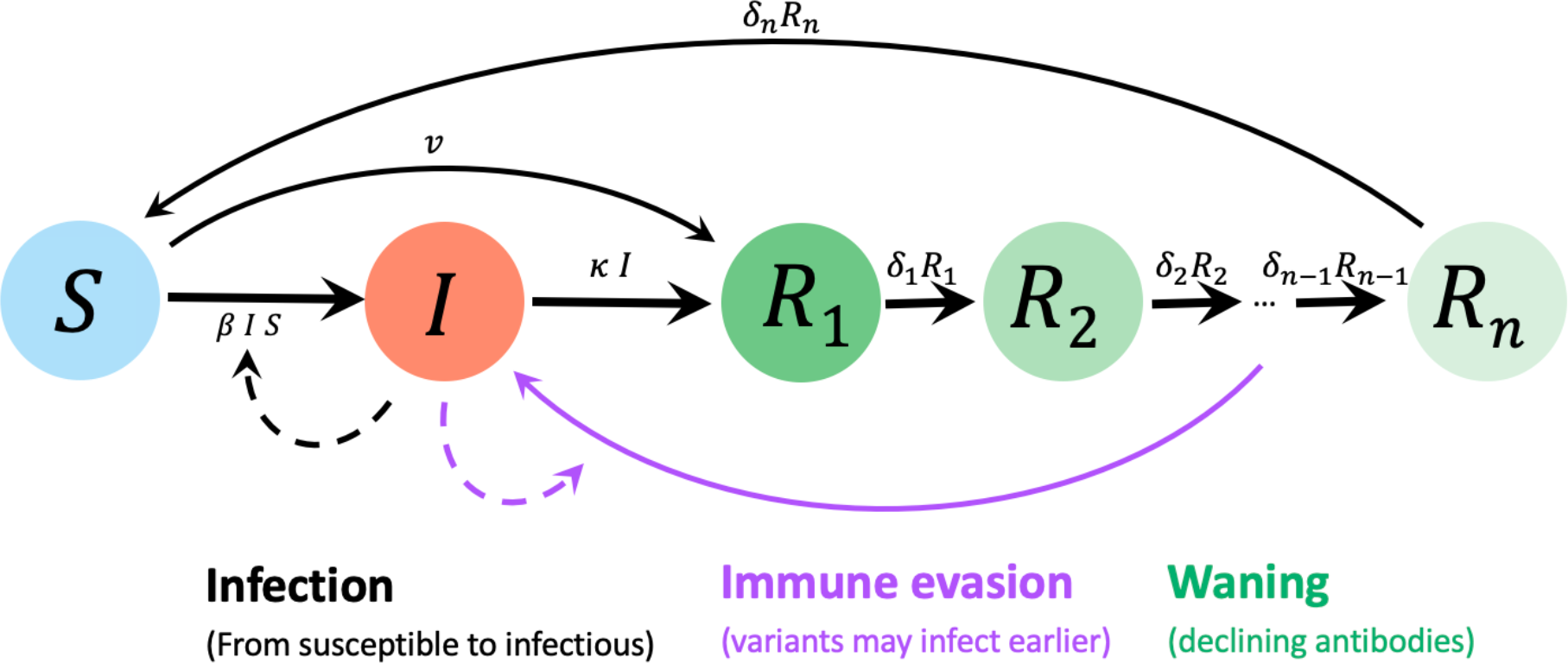
Epidemiological model used to predict impact of changing variants, behaviour, and policy on endemic levels of disease. We consider populations that have a high level of immunity due to prior infection and/or vaccination and that consist of *S*: a susceptible fraction, *I*: an infected fraction, and *R*_*j*_: a recovered fraction with immunity at different stages of waning. Parameters are *β*: transmission rate, *κ*: recovery rate, *δ*_!_: per-class waning rate per day, and *v*: vaccination rate at the population level, all measured in the present-day population with prior exposure. Movement between adjacent recovered classes is set equal to *δ*_!_ = *nδ*, so that the expected time between first recovering and returning to the susceptible state is 1/*δ* days.

As we are modelling the long-term epidemiological dynamics in a population previously exposed to the virus via vaccinations and/or infections, we emphasize that the susceptible class, *S*, consists of individuals who have had previous exposure but are currently susceptible due to waning immunity. Throughout this paper, we are thus describing the epidemiological dynamics in a previously challenged population.

When we model vaccination, vaccines move individuals from the susceptible (*S*) to the first recovered class (*R*_1_) at rate *v* per population (Figure 2). We consider *v* to be the total fraction of the population moved (rather than the rate per susceptible individual) to align with data on observed or target vaccination rates within a country. Vaccination is therefore protective against infection in this model until vaccine-induced immunity wanes, which occurs at the same rate that infection-induced immunity wanes (although we do extend the model to consider the possibility that vaccination does not elicit an immune reaction in some individuals). We do not separately model disease or severity, or vaccine’s effectiveness against these as distinct from protection against infection.

Before considering variants or NPI measures, the dynamics describing changes in the number of individuals in each compartment within the SIR_n_ model are:

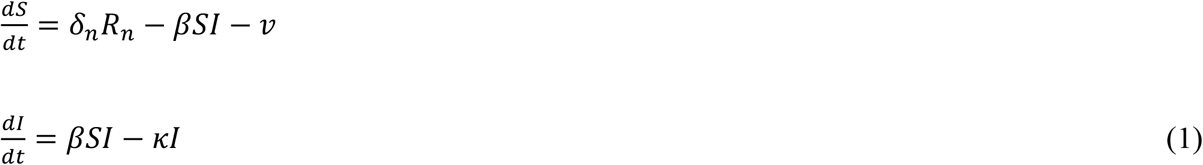

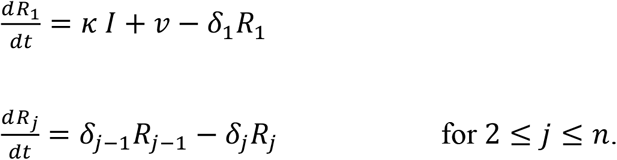

We can find the equilibria of this system of equations by setting the derivatives to zero and solving, yielding two equilibria for the fraction of individuals in each class. One equilibrium corresponds to the disease being absent (*Ŝ* = 1), and the other to the disease being endemic:

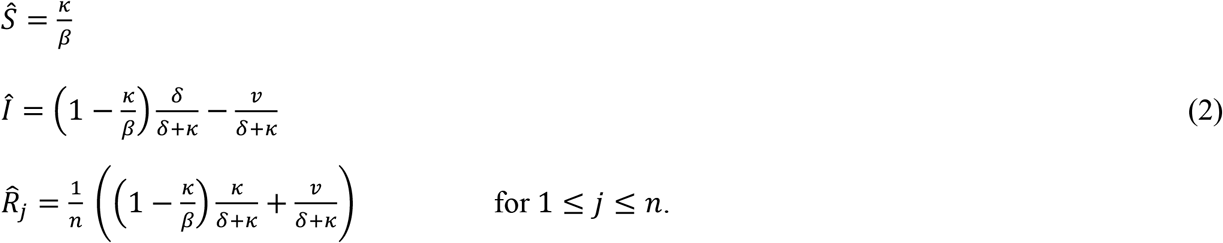

Importantly, because we are explicitly modelling endemic COVID-19 most individuals have previously been exposed to SARS-CoV-2, susceptibility and infectiousness may be lower in the current population than when the virus first appeared in humans because of cellular immunity, any residual humoral immunity among susceptible individuals (Tan et al. 2023), and/or due to any behavioural changes (including better ventilation, testing and self-isolation practices). For example, the rapid induction of cellular immunity reduces the viral load of typical breakthrough infections (Puhach et al. 2022), lowering transmission (*β*) compared to a fully naïve population. The epidemiological dynamics in this endemic model thus depend on an “*endemic basic reproductive number*”, 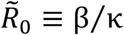, which is the basic reproductive number in a population consisting entirely of currently susceptible, but previously vaccinated or infected, individuals whose immunity has waned. In contrast to the initial *R*_0_ for COVID-19 at the time of its emergence, the parameters of the endemic model and in 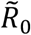 (transmission, β, and recovery, κ) refer to rates in this previously challenged population. Our estimates of 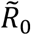 range from 1 to 6 (Appendix 1), depending on estimates used for recovery rates, waning, and the endemic level of infections within a population, with a value of 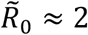 2 for the parameters considered typical (Table S1).

If this previously challenged population were fully susceptible (*Ŝ* near one), the disease would spread when rare as long as transmission rates were higher than recovery rates, 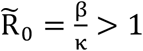, which we assume to hold. In this case, the endemic equilibrium (2) exists and is stable for all examples considered here. The endemic equilibrium may, however, be unstable (Hethcote, Stech, and Van Den Driessche 1981), leading to sustained cyclic dynamics, outside of the parameters used here (e.g., for *n* large enough).

The equilibrium can also be written in terms of 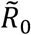 as:

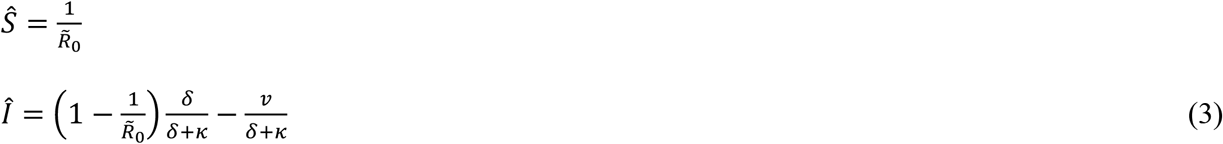

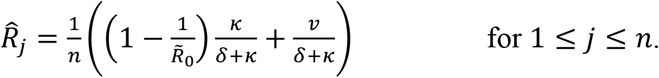

Also of relevance is the number of bouts of disease that an individual expects per year, which is 365 *β Ŝ Î* = 365 *κ Î*, assuming average behaviour (see Table S1).

Given that recovery rates are higher than waning rates (*δ*≪ *κ*), equation (3) shows that the number of infectious individuals at the endemic equilibrium is reduced by the number of vaccinations within a typical recovery period (*v*/*κ*). If more vaccinations were to be given in a typical recovery period than the fraction of individuals expected to be infectious in the absence of vaccination, the disease could be driven extinct locally (though we note that in this model vaccination has a very high, if temporary, efficacy against infection, and that reintroductions are expected from importations, animal reservoirs, and chronic infections). Uptake of additional vaccine doses during 2023 has, however, been so low in many countries as to make little difference to the incidence and dynamics of SARS-CoV-2 (e.g., daily [annual] rates of 0.012% [4.7%] in France and 0.023% [8.8%] in the United States from 1 January April, 2023; (Our World in Data 2023)). To simplify the discussion, we ignore ongoing vaccination for now, returning later to a discussion of the impact that vaccination uptake can have on individual risks of infection and on the overall incidence of disease.

### Spread of variants during the endemic phase

A new variant may spread within the population if it is more transmissible (e.g., better binding to ACE2 receptors on host cells), more immune evasive, or both (see (Cao et al. 2023) for empirical measures for SARS-CoV-2). We can calculate the rate of spread of a variant using the SIR_n_ model by allowing different transmission rates for the resident variant (*β*) and the new variant (*β*^∗^ = *β* + ∆*β*) and by allowing immune evasive variants to infect earlier than the resident strain, while antibody levels are at intermediate levels. Specifically, we assume that an immune evasive variant can infect the last *m* recovered classes (each of which is at frequency 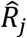 at the endemic equilibrium given by (3)), as well as susceptible individuals.

As described in Appendix 1, a new variant introduced into a population at the endemic equilibrium has a selective advantage of:

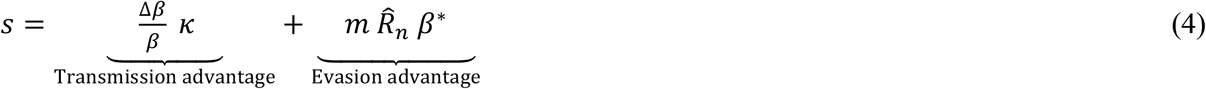

The selection coefficient, *s*, describes the rate at which the new variant spreads relative to the resident variant. Selection coefficients describing evolutionary changes in SARS-CoV-2 have been estimated in many jurisdictions using sequence information and are often relatively stable over time and space when measured consistently against the same reference strain (van Dorp et al. 2021; Otto et al. 2021).

What are the consequences of spreading variants for the incidence of disease? The incidence is initially expected to rise exponentially at rate proportional to selection (specifically, *s Î*), but this is only transient as the new variant spreads through the susceptible population available to it. Over the long term, we show that the impact on the endemic level of disease depends strongly on whether the variant increases transmission rates and/or increases immune evasiveness, as well as the persistence of immune evasion during subsequent infections, even for variants with the same selective advantage.

In particular, when the population is comprised entirely of the new variant, the endemic level of disease changes to:

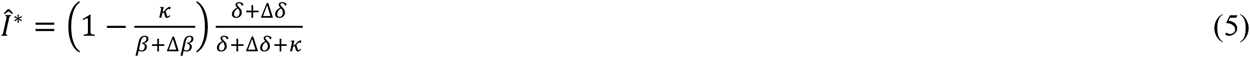

(found by solving equation (A1) for the endemic equilibrium when only the variant is present). The term ∆*δ* refers to how the variant changes the rate of complete waning, from first entering the recovered class to returning to the susceptible state (i.e., 1/(*δ*+ ∆*δ*) is the mean number of days to return to susceptibility). [As short-hand, we refer to a variant’s impact on immune evasion as a change in the waning rate ∆*δ*, but the model actually assumes log-antibody levels wane at a constant rate but the variants can just infect earlier, becoming susceptible sooner.]

Equation (5) allows us to evaluate the long-term impact of different types of variants. For immune evasive variants, the results are strongly dependent on the variant-specific immunity that develops after infection, even if the lineages have the same selective advantage and rate of spread (*s*, equation (4)). Consider two extreme possibilities:

- **Transient immune evasiveness:** If the variant better evades the initial suite of antibodies but causes infections that generate variant-specific immunity, subsequent infections may no longer be immune evasive. In this case, subsequent infections would require the full waning period (returning to the *S* compartment and not the *R*_*i*_ compartments), as for the resident strain. With only transient evasiveness, the long-term level of COVID-19 is ***unaffected*** (∆*δ*= 0).
- **Persistent immune evasiveness:** If the variant allows infections to occur earlier, both in the first and in subsequent infections, then the rate of return to susceptibility is consistently higher 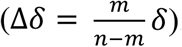. With waning slow relative to recovery (*δ*+ ∆*δ*≪ *κ*), persistently immune evasive variants cause the incidence of disease to ***rise in proportion*** to the increased rate of waning, *Î* ^∗^ ≈ *Î* (1 + ∆*δ*/*δ*).

In either case, immune-evasive variants spread in the short-term because of the selective advantage, *s*, gained by infecting susceptible individuals earlier, but the subsequent dynamics and long-term impact differ greatly, depending on whether variant-specific immunity builds (Figure 3). The extent to which exposure to a variant elicits variant-specific humoral or cellular immunity almost certainly falls between these two extremes. Metanalyses suggest, for example, that infections with Omicron are less protective against reinfection with Omicron, compared to the protective effects of pre-Omicron variants, suggesting some persistence in its immune-evasive properties (Arabi et al. 2023).

**FIGURE 3:**
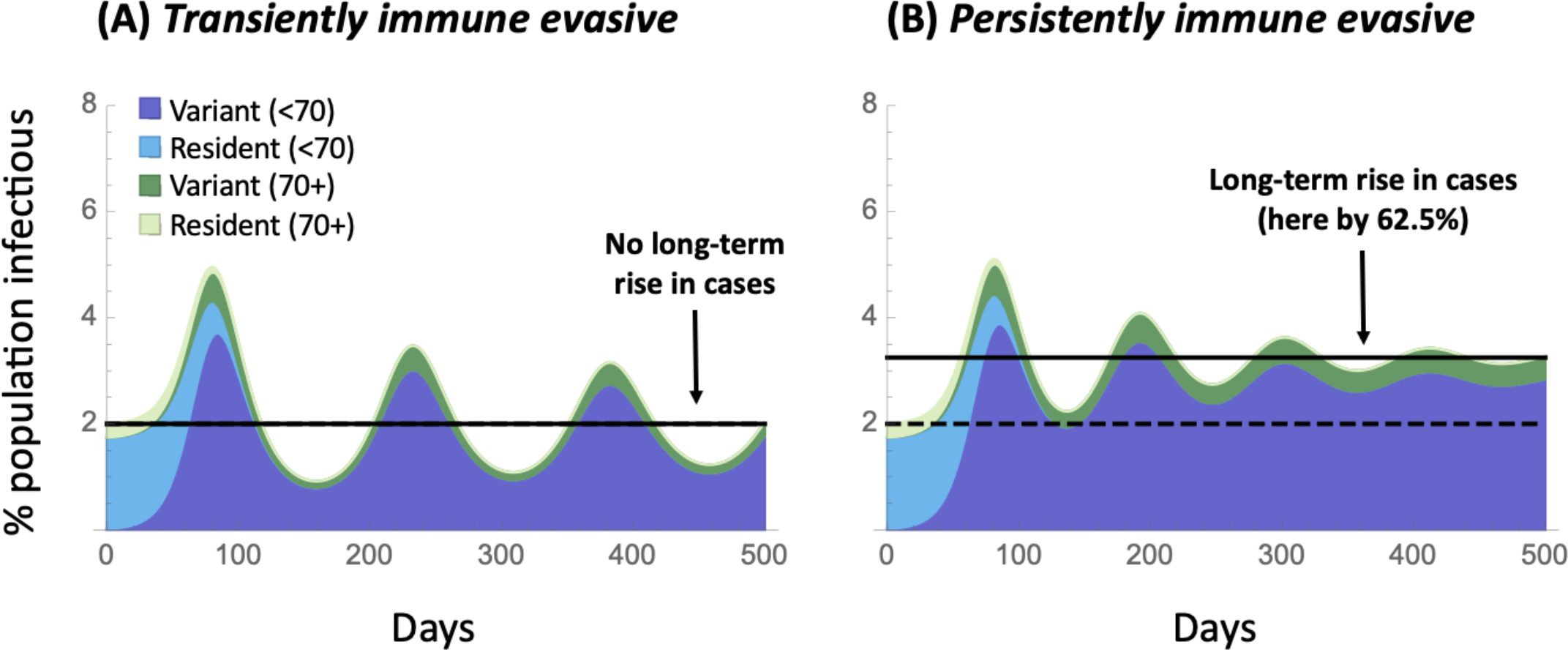
Impact of the spread of immune evasive variants depends on whether a variant-specific immune response is elicited. Plots illustrate the dynamics over time of a more immune evasive variant, which is able to infect earlier in the waning period (by *m* = 2 out of *n* = 5 recovered classes), giving the variant an *s* = 8.3% selective advantage per day, which lies in the range of the faster spreading variants observed in the past year (CoVaRR-Net’s CAMEO 2023). While the short-term spread of the variant (dark shading taking over from light shading) and rise in cases are nearly identical (given *s* is the same), the long-term consequences differ substantially depending on whether the variant’s evasive properties are (panel A) transient or (panel B) persistent. The endemic equilibrium rises only if evasiveness persists in subsequent infections (panel B). We illustrate the dynamics in younger (under 70) and older (70+) individuals, who are more prone to severe cases. Parameters: *κ* = 0.2, *δ*= 0.008, *Î* = 2%, *β* = 0.42, the nominal parameter estimates given in Appendix 1 for all age classes.

By contrast, more transmissible variants change the endemic equilibrium to:

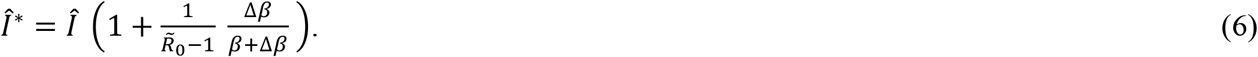

As a consequence:

- **More transmissible variants:** If the variant increases transmission rate, spread in the short term depends directly on the change in transmission (*s* = (∆*β*/*β*) *κ*; equation (4)), while the long-term impact on the incidence of disease exhibits diminishing returns (*Î* ^∗^/*Î* depends on ∆*β*/(*β* + ∆*β*)). Thus, more transmissible variants have **a less than proportionate influence** on the number of cases in the long term, unless the susceptible pool is large and 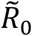 small 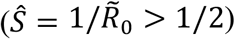.

While more immune evasive variants increase the pool of susceptible individuals available to them (in our model, by adding the last *m* recovered classes to the susceptible class), more transmissible variants deplete the susceptible pool. This can be seen by the effect on the susceptible class at equilibrium, which decreases from *Ŝ* = *κ*/*β* for the resident virus to *Ŝ* ^∗^ = *κ*/(*β* + ∆*β*) for a more transmissible virus. For this reason, more transmissible viruses are, to some extent, self-limiting and often have less of an effect on the long-term number of cases than seen for a permanently immune evasive variant. Thus is illustrated in Figure 4, which shows that the equilibrium rises less for a given % increase in transmissibility (panel C) than for the same % increase in waning rate for a permanently immune evasive variant (panel B), unless 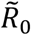 is so small that most individuals in the population are susceptible. Of course, variants may combine features affecting transmissibility and immune evasiveness (Cao et al. 2023), as explored in Figure S1.

**FIGURE 4:**
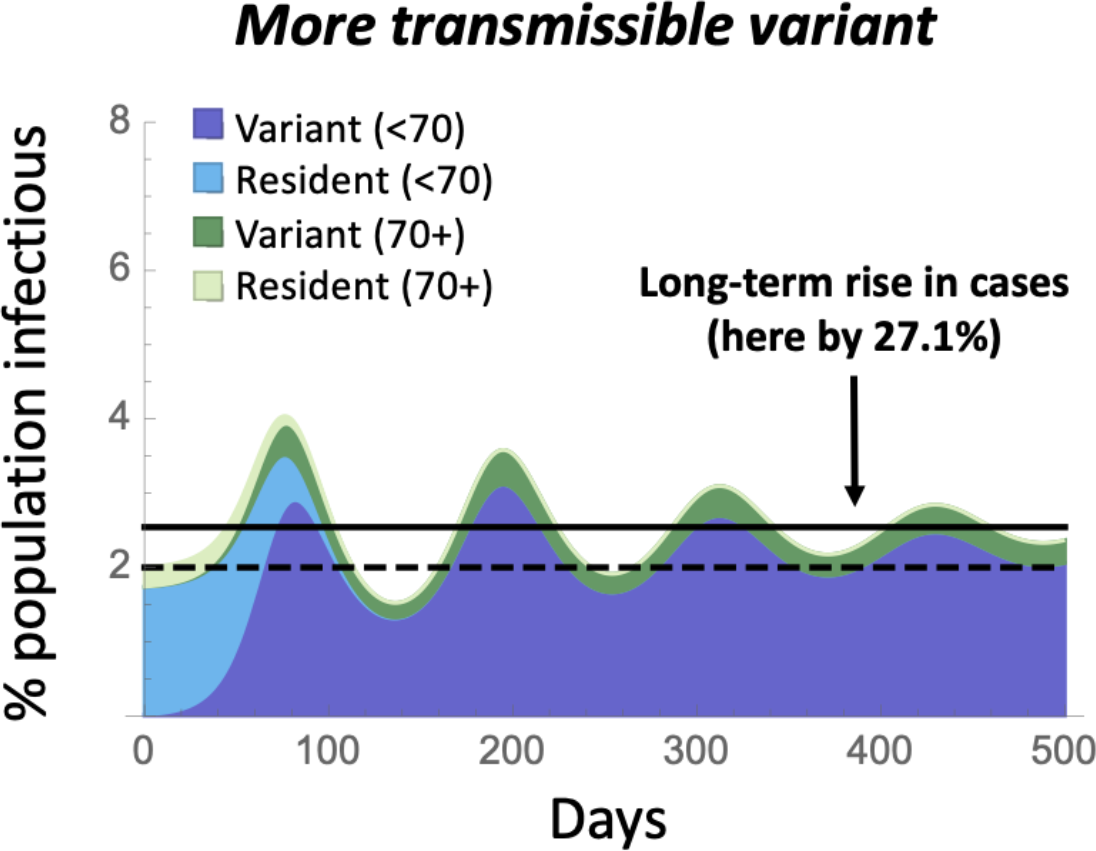
Impact of the spread of a more transmissible variant. Plot illustrates the dynamics over time of a more transmissible variant, which increases *β* (and hence 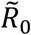) by 42%, chosen to give the variant the same selective advantage as in Figure 3 (*s* = 8.3%). While exhibiting a similar short-term rise in cases as in Figure 3, the long-term impact is intermediate. Parameters are identical to Figure 3, with *β*^∗^ = 0.59.

This model predicts, as seen in the data over the past year (Figure 1), only modest rises and falls in case numbers, despite substantial evolutionary change in the frequency of different variants. This occurs because of the substantial population-level immunity that persists in the population at the endemic equilibrium. With a mixture of susceptible, infectious, and recovered individuals, the effective reproductive number at the endemic equilibrium must be one (*R*_*e,t*_ = 1) for the resident and *R*_*e,t*_ = 1 + ∆*β*/*β* for a more transmissible variant (e.g., *R*_*e,t*_ = 1.42 in Figure 4). Thus, only a modest drop in the susceptible population is needed (1/*R*_*e,t*_) before the infectious class peaks and falls again.

The robustness of these conclusions is explored in Appendix 2, considering different models of immunity (including leaky immunity) and the inclusion of features such as an exposed class and failure to seroconvert. The strength of selection (equation (4)) and the long-term impact on endemic levels of disease (equation (5)) are robustly observed. The speed at which the waves dissipate over time, however, is sensitive to model assumptions, stabilizing faster than observed above in many cases (Figure S2), so we caution that the nature of subsequent wavelets caused by the spread of a variant is hard to predict.

We conclude that variants may have dramatically different long-term impacts on the level of disease depending on the nature of the advantage (transiently or persistently immune evasive and/or more transmissible), despite exhibiting the same selective advantage and hence spreading at the same rate (e.g., with selection of *s* = 8.3% per day in Figures 3 and 4). Indeed, a variant that is transiently immune evasive but less transmissible can spread and would be expected to reduce the equilibrium level of disease, except that once immunity to this variant has built, the previous resident reemerges because of its higher transmissibility (Figure S1B).

Figure 5 shows these long-term impacts on disease incidence at the endemic equilibrium across the range of plausible parameters (Appendix 1). Transiently immune evasive variants have no long-term impact (panel A), whereas the rise in cases is nearly proportional to the ability of a variant to evade immunity, if that evasiveness is persistent, regardless of the exact parameter values (panel B). By contrast, the long-term impact of a more transmissible variant depends strongly on the current transmissibility, as measured by the endemic reproductive number. The larger 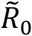 is, the smaller the long-term impact of more transmissible variants is on disease levels (Figure 5C), essentially because the pool of susceptible individuals is then smaller and rapidly depleted by more transmissible variants 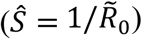. That said, for given waning (*δ*) and recovery (*κ*) rates, the endemic level of disease is higher when 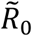 is higher (equation (3)), so a small percentage increase in disease incidence can still have a numerically important impact on the burden of disease.

**FIGURE 5:**
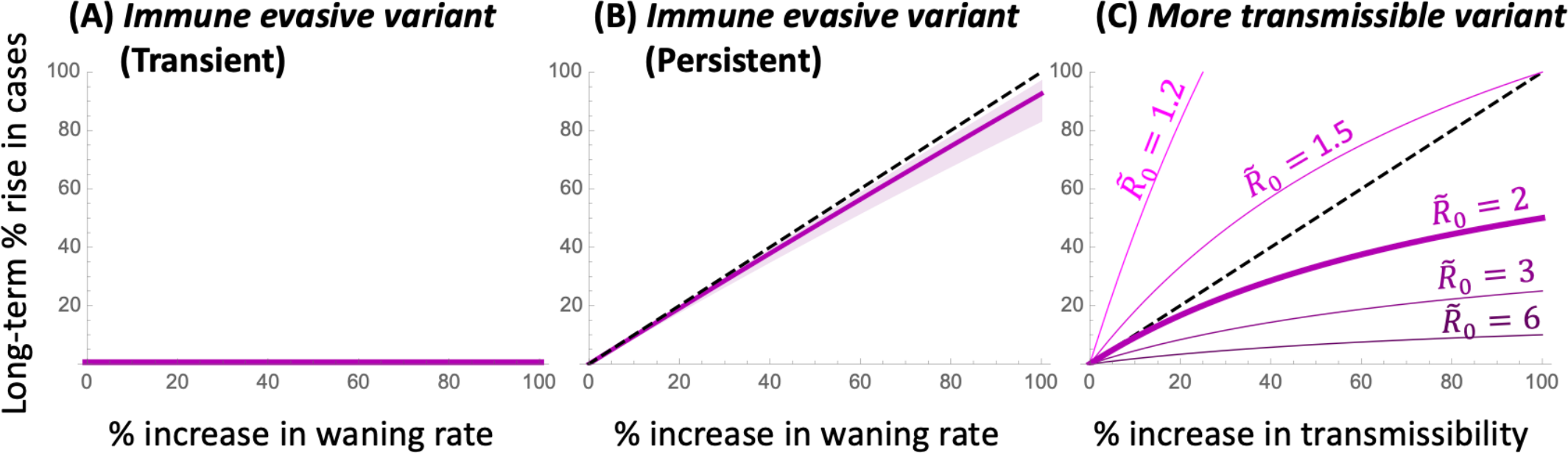
Long-term impact of a variant. The percent change in the endemic equilibrium is shown as a function of the percent by which the variant increases the rate at which recovered individuals become susceptible again (∆*δ*, panels A,B) or transmissibility (∆*β*, panel C). Transiently immune evasive variants have no long-term impact, while persistently immune evasive variants cause the endemic incidence of disease to rise nearly in proportion (dashed curve) across all parameters considered plausible (purple shading, with the nominal parameter values illustrated by a thick purple curve; see Appendix 1 and Table S1). By contrast, the impact of a more transmissible variant that increases *β* depends strongly on 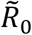 (but none of the other parameters), leading to a less than proportional rise in cases whenever 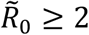 (see equation (6)).

### Controlling an endemic disease

In the face of variants that are increasingly transmissible and/or persistently immune evasive, the endemic level of disease is expected to rise over time, but these increases can be countered by protective measures at the individual and population level. Protective measures range from vaccination to NPI measures, such as testing and self-isolation, avoiding crowded indoor spaces, improving ventilation, and the wearing of well-fitting and high-quality masks (The Royal Society 2023). Here we explore the impact of these protective measures at both the individual level, modulating the frequency of infections, and the population level, modulating the endemic incidence of disease.

#### Vaccination

Vaccination allows individuals to short-circuit the disease cycle by boosting antibody levels and immunity by another dose rather than by infection. Globally, 65% of people have had the primary series of COVID-19 vaccines but an average of only 0.35 booster doses have been distributed per person ((Our World in Data 2023); accessed 22 August 2023). Jurisdictions vary widely in recommended vaccine schedules and access to vaccines. For example, only individuals at higher risk of serious illness are eligible for COVID-19 vaccinations in the United Kingdom (NHS 2023). In Canada, the National Advisory Committee on Immunization recommends that all adults be offered vaccines six months after the last dose or infection (NACI 2023). The Centre for Disease Control in the USA, however, recommends that individuals stay up to date with important vaccine updates (e.g., the updated mRNA vaccines providing protection against BA.4 and BA.5 in the fall of 2022 and against XBB in the fall of 2023 (CDC 2023)).

There is substantial uncertainty and confusion in both public and public health circles about the value of regular vaccinations against COVID-19 (Lin et al. 2023). Here, we explore one aspect: how much do regular vaccinations reduce the burden of disease expected at an endemic equilibrium?

We consider the impact of policies aimed at future vaccine uptake, encouraging vaccination of a portion of the population (*v*) per day. Given that vaccines are recommend only after a substantial amount of time has passed since the previous dose or infection, these vaccinees are assumed to target individuals in the susceptible class, moving them into the first recovered class (*S* to *R*_1_; equation (1)). Unlike many previous epidemiological models (reviewed in (Scherer and McLean 2002), we assume that the target vaccination rate is set by policy, adjusting public health campaigns, vaccine cost, and availability to meet these targets (i.e., *dS*/*dt* in equation (1) declines by a fixed daily rate, *v*, rather than a per capita rate, *v S*).

We consider vaccination rates in Canada as typical of what can be achieved when vaccines are available at regular intervals (every six months). From April-July 2023, vaccination rates in Canada have averaged only 0.012% of the population per day (annual rate of 4.5% (Health Infobase Canada 2023)). While these vaccinations help those individuals receiving a dose, this level has a negligible impact on the endemic level of cases (decreasing *Î* from 2% to 1.94% for the nominal parameter values). Many public health agencies have encouraged COVID-19 vaccine updates in the fall (Mahase 2023). For example, vaccination rates in Canada during September-December 2022 were 14 times higher (0.174% of the population per day, an annual rate of 63.5% (Health Infobase Canada 2023)), a rate that substantially lowers the endemic equilibrium level if maintained (from 2% to 1.16% for the nominal parameter values).

At an individual level, vaccination reduces the number of infections that one expects to have. Individuals on a regular six-month vaccination schedule are expected to be protected from neutralizing antibodies for 1/*δ*out of every 180 days. Calculating the probability of waning and infection before their next vaccine (equation (A2); Appendix 1), a regularly vaccinated individual expects to have about 60% as many infections per year (0.88 vs 1.46) for the nominal parameter values (Appendix 1). Across the range of parameters considered plausible, vaccination every six months leads to only 40%-66% as many infections annually (supplementary *Mathematica* file).

At a population level, in our model, vaccination reduces the endemic level of infections to 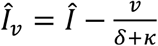. (equation (2)). That is, the endemic level of infections is reduced by approximately the number of vaccinations conducted during the infectious period (*v*/*κ*, given that waning is considerably slower than recovery). Figure 6 illustrates the impact of increasing and maintaining the vaccination rate at the higher levels observed in Canada in the fall of 2022 (*v* = 0.174%). In this case, the long-term incidence of infection can be driven down by ∼42% (panel A). Across the range of parameters considered in Appendix 1, this population-level benefit ranging from a 12-100% decline in incidence of disease, falling at the lower end of the benefit when disease incidence is high without vaccination (*Î* = 4%) but at the higher end and allowing complete eradication when disease incidence is low without vaccination (*Î* = 0.5%).

**FIGURE 6:**
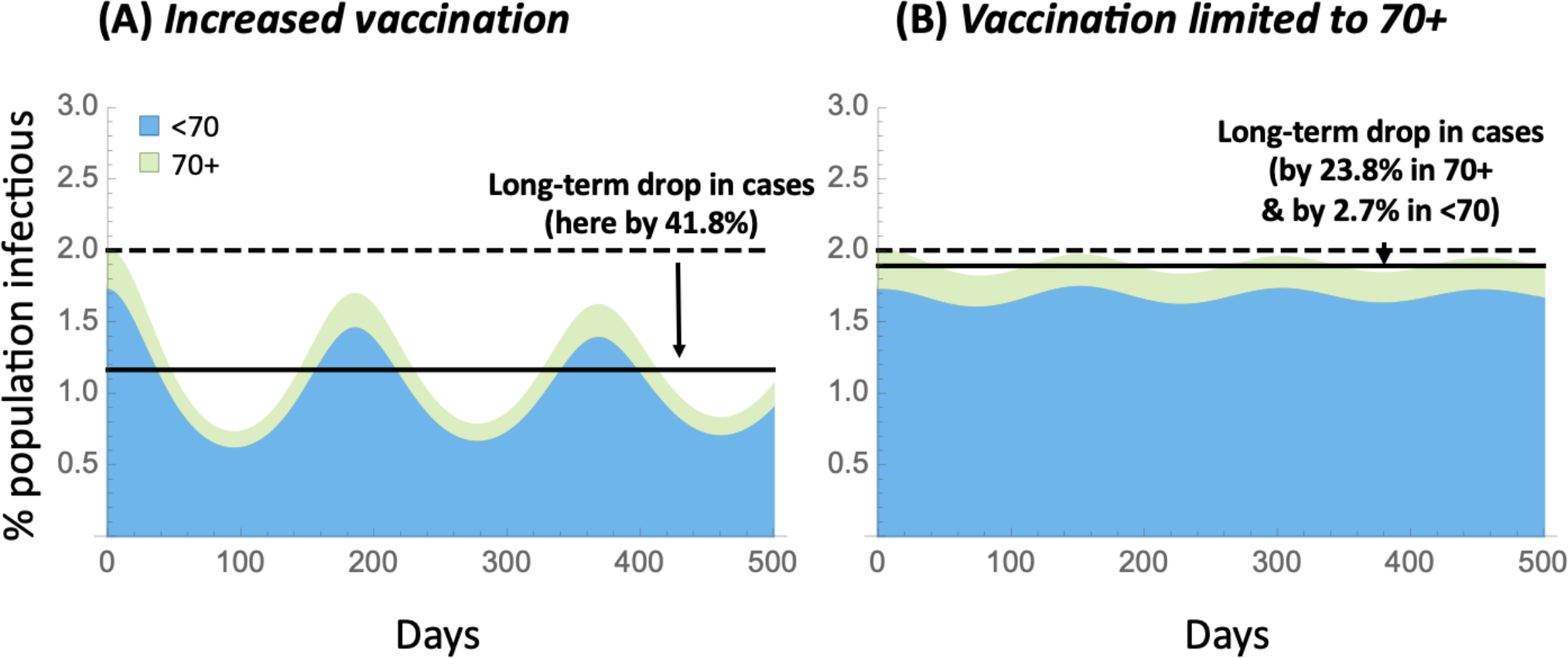
Impact of vaccination strategies. Plots illustrate the dynamics over time when vaccination is increased to 0.174% of the population per day (annual rate of 63.5%), either (A) within the entire population or (B) limited to a more vulnerable population (illustrated as 70+ in age). Parameters are identical to Figure 3, with *v* = 0.00174.

One policy option considered in many jurisdictions is to regularly vaccinate only the more vulnerable segment of the population. Figure 6B illustrates, however, that limiting vaccination to the more vulnerable population (shown here as vaccinating only those over 70 at a rate *v* = 0.174%) has less impact on the frequency of infections experienced by this vulnerable population (reduced by only 24%), because the incidence of COVID-19 remains high overall, increasing their risk of exposure.

That said, the additional protection provided by COVID-19 vaccines against severe disease, above and beyond the protection provided against infection, means that the risk of hospitalization and death can be lowered by vaccinating the vulnerable (Nyberg et al. 2022; Chemaitelly et al. 2022). Further reducing the risk of infection and severe disease, however, requires a broader vaccination campaign. Broad vaccination campaigns provide additional protection for the vulnerable, while also reducing the number of sick days, risks of long COVID, and severe disease among those not known to be vulnerable.

As noted in Appendix 2, seroconversion rates upon vaccination are high (Wei et al. 2021), so we do not correct *v* for the small fraction of doses that fail to elicit an immune response. Not all individuals will, however, achieve high levels of immunity following vaccination and not all will be protected from infection. A mixture of induced immunity could be modelled by moving vaccinated individuals into a distribution of *R*_!_ classes. Additionally, some individuals being vaccinated may have been exposed in the recent past (i.e., coming from the infectious or recovered compartments, not solely from the susceptible classes). These possibilities are not explicitly modeled, although they would lower the protection offered by vaccination, akin to lowering *v*, and so require higher vaccination uptake to achieve the benefits described above.

### NPI measures

A wide variety of non-pharmaceutical interventions have been deployed to counter the spread of SARS-CoV-2, including testing and self-isolation, enhancing ventilation and air filtration, and wearing of high quality masks (see evaluation of evidence in the report (The Royal Society 2023)). Here, we consider the individual-level and population-level benefits of NPIs, as a function of their impact on preventing transmission of the virus, modelled by NPIs preventing a portion *p* of transmissions both from and to NPI users. Specifically, we assume that the NPI measures reduce transmission from *β* to (1 −*p*) *β* if one member in an interaction practices the measures and to (1 −*p*)^2^ *β* if both do. The population is assumed to be heterogenous, as illustrated in Figure S4, consisting of a fraction *f* who regularly engage in NPI measures and are denoted by a ‡ (compartments *S*^‡^, *I*^‡^, 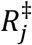 for *j* = 1 to *n*, which sum to *f*) and a fraction 1 −*f* who do not (compartments *S, I, R*_*j*_, which sum to 1 −*f*). See Appendix 3 for model details.

We first determine the benefit to an individual who adheres to NPI measures (e.g., masking). At the endemic equilibrium, an individual engaging in the NPI measure has a lower risk of being infected at any given point in time of:

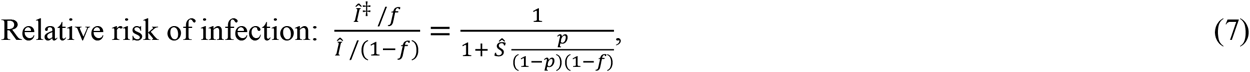

where *Ŝ* is the fraction of the population at the endemic equilibrium who are susceptible and do not engage in the NPI measure (given by (A11), *Ŝ* ≈ *κ*/*β* when *f* is small). This relative risk is mathematically equivalent to the relative rate at which individuals become infected for those who do versus do not engage in the NPI measure, as well as the relative number of infections expected per year.

Figure 7 illustrates the relative risk of infection (equation (7)). As expected, the more effective the NPI measure is (the higher *p*) the lower the relative risk to individuals who engage in the NPIs (panel A). This individual-level benefit is only weakly dependent on the fraction of the population currently engaging in these measures (*f* = 10%, 50%, and 90% shown as solid, dashed, and dotted curves), with the relative risk rising slightly as *f* increases because non-practitioners gain a slight benefit from those who do practice the NPI measure.

**FIGURE 7:**
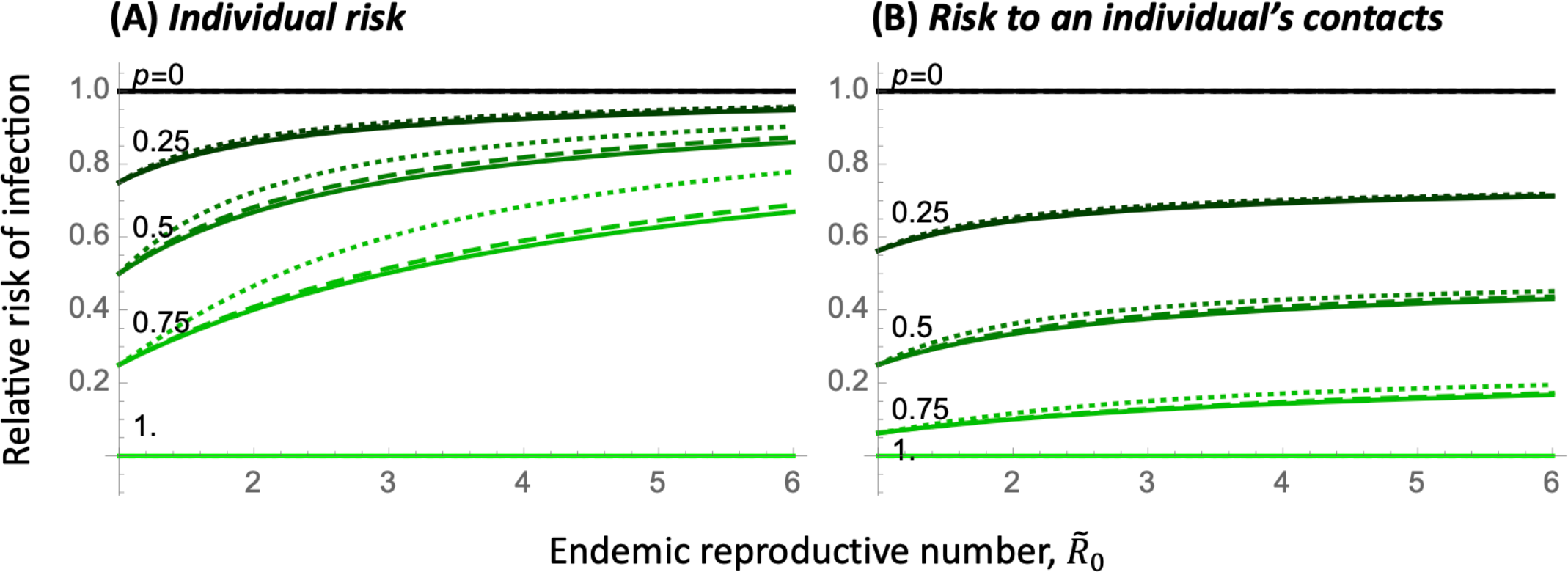
Risk of infection for individuals regularly engaging in an NPI measure such as masking, relative to unmasked individuals. Coloured lines illustrate different levels of protection, *p*, provided by the NPI measure, in a population where the fraction of individuals engaging in the NPI measure is *f* = 10% (solid), 50% (dashed), and 90% (dotted). Panel A shows the risk of infection and panel B the risk of becoming infected and infecting a contact for an individual engaging in NPI measures, relative to those who do not. The x-axis gives the endemic reproductive number in this heterogeneous population, 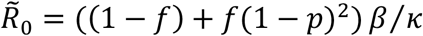. Parameters: Relative risk depends on the parameters only through 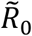, *f*, and *p*.

The individual benefits depend strongly, however, on the endemic reproductive number of the disease 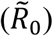 At the nominal value of 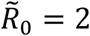 and assuming low population-level uptake (*f* small), a person’s relative risk of infection can be substantially reduced for NPIs that provide fairly modest protection (by 14% and by 33% for *p* = 25% and 50%, respectively). For 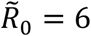 (on the high end of the range considered plausible; Appendix 1), however, these individual-level benefits diminish (to 5% and 14% for *p* = 25% and 50%, respectively), because individuals are exposed so often that modestly protective NPIs only moderately delay infection.

Not only is a susceptible individual who regularly engages in NPI measures less likely to become infected when in contact with an infectious individual (by a factor 1 −*p*), but they are also less likely to pass the infection on to one of their contacts (by another factor 1 −*p*), compared to a non-practicing individual who is currently susceptible. Accounting for the proportion of time that practicing and non-practicing individuals are susceptible (as in equation (7)), the risk per unit time of being infectious and infecting a contact is substantially lowered for those engaging in NPI measures relative to those who do not (Figure 7B). This validates the approach used by many who adhere to NPI measures, such as masking, in order to protect vulnerable relatives and close contacts.

The curvature of the relative risks in Figure 7 highlights the utility of multiple complementary interventions: other policies that reduce transmission (lowering _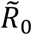_) make masking more effective to individuals, because those individuals are less repeatedly exposed.

The individual-level benefits of NPI measures diminish with 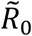 (x-axis of Figure 7) because individuals practicing NPI measures are more likely than non-practitioners to have remained uninfected and so are more often susceptible at the time of exposure, which increases their relative risk of infection as 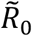 increases. Different results are obtained if individuals frequently switch their behaviour (e.g., masking some days and not others). Modifying the model as described by equation (A12), all individuals are then equally likely to be susceptible on any given day, and the NPI measure always reduces the risk of infection by a factor (1 −*p*) for each practicing individual in an interaction. That is, the benefits remain at their maximal value of (1 −*p*) in panel A and (1 −*p*)^2^ in panel B, regardless of 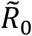 and *f* (Appendix 3).

We next evaluate the population-level advantages of NPI measures by calculating the fraction of infected individuals expected at the endemic equilibrium (*Î* + *Î* ^‡^) when a fraction *f* of the population upholds these measures, relative to a population in which nobody does:

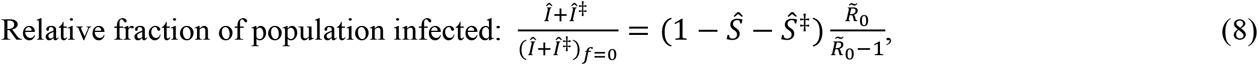

using the equilibrium values given by equation (A11). The right-hand side of equation (8) emphasizes that the population-wide benefits increase (fewer people will be infected) when there are more susceptible individuals available (*Ŝ* + *Ŝ* ^‡^ larger).

While the population-level impact is small when few individuals mask (left panel of Figure 8), there are substantial benefits to having moderate to high adherence to the NPI measures (central and right panel). These benefits are strongest when the endemic reproductive number is small, potentially moving the population away from the endemic case, where COVID-19 persists, to the disease-free equilibrium (when the curves cross the x-axis), again emphasizing the added benefits that come from combining interventions.

**FIGURE 8:**
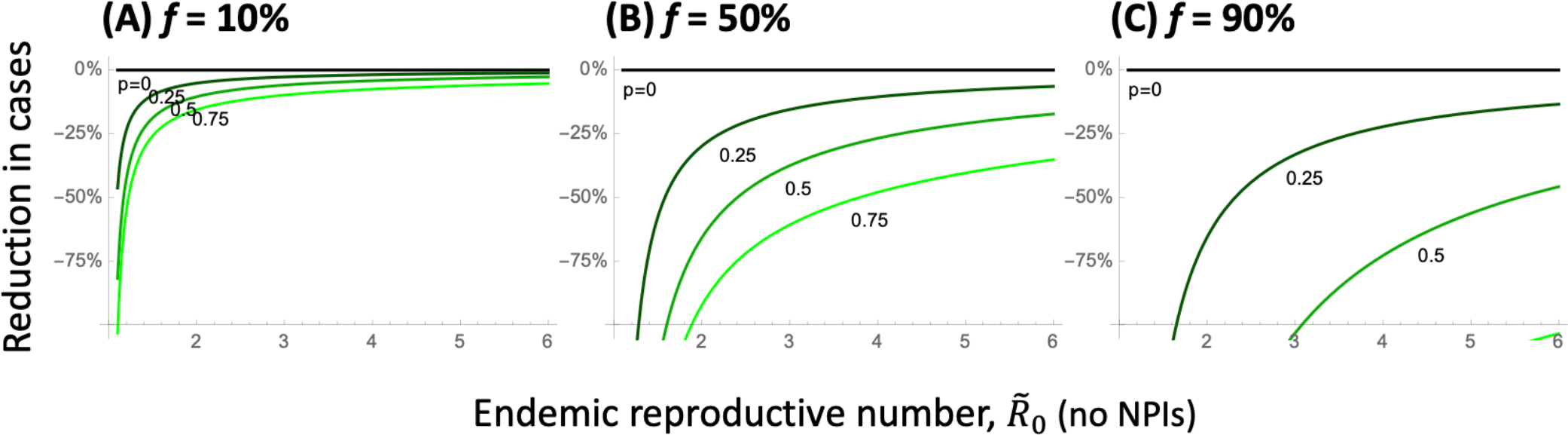
Reduction in cases at the endemic equilibrium when a fraction *f* of the population engages in an NPI measure, relative to when none do. Coloured curves illustrate different levels of protection, *p*, provided by the NPI measure. Panels show the fraction of the population practicing the NPI measure: (A) *f* = 10%, (B) 50%, and (C) 90%. The x-axis gives the endemic reproductive number in a population that is not engaging in the NPI measure, given by 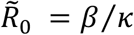. Parameters: Reduction in cases depends on the parameters only through 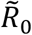, *f*, and *p*.

The reduction in cases caused by NPI measures is expected to result in a proportionate reduction in severe cases and deaths. Even a modest reduction (say 20%) can have non-linear benefits when hospitals are at capacity, improving care for all (Wichmann and Wichmann 2023). Achieving such benefits at a population level, however, requires that there be clear messaging and incentives to obtain the moderate to high levels of uptake required to impact population-wide infection rates.

## Discussion

This paper aims to expand our understanding of the impact of variants, as well as behavioural and public health measures, on endemic diseases like COVID-19. Widespread measures, both by individuals and public health agencies, repeatedly “flattened the curve” of COVID-19 during the first two years of the pandemic, reducing viral transmission to save lives and avoid collapse of health care systems (Ogden et al. 2022; Talic et al. 2021). Since mid-2022, however, COVID-19 has persisted at high levels throughout the world, becoming endemic with no sign of abating even during summer months. The mantra to “flatten the curve” is no longer relevant, as endemic levels are already fairly flat, and we lack a compelling guide to govern our collective behaviour in its place.

For COVID-19, endemic does not mean constant, with wavelets caused by variants, changing behaviour, and varying vaccination rates. Nor does endemic mean rare, as on-going high levels of COVID-19 health impacts remain. Nor does endemic mean out of our control, as protective measures continue to have important benefits, boosting immunity through vaccination and reducing transmission through effective NPI measures. The goal of this paper is two-fold: to explore the impact of evolutionary changes in the virus on disease incidence and to discuss how protective measures can counteract these rises, reducing disease risks.

Variants of endemic diseases that increase transmissibility and/or immune evasion are selectively favoured, with rises in frequency that can be measured empirically, yielding estimates of the strength of selection (*s*). While the strength of selection accurately predicts the speed with which one variant replaces another, it does not predict the long-term impact on endemic levels of disease. For a given selection coefficient, we have shown that the long-term impact on disease is negligible for variants that are more immune evasive, but only transiently so, eliciting variant-specific antibodies that protect from reinfection (Figure 5A). By contrast, immune evasive variants that fail to elicit variant-specific antibodies have a persistent advantage, leading to a nearly proportional increase in cases in the long term (Figure 5B). In Appendix 2, we also consider variants that cause immunity to become leakier, increasing the risk of infection for all recovered classes, which are particularly problematic (Figure S3), causing a high long-term rise in cases because all individuals remain prone to infection if leakiness is persistent. Variants that are more transmissible generally have an intermediate impact on disease incidence (Figure 5C). Thus, depending on the exact properties of new variants, we may see smaller or larger rises in cases over the long term, even for variants initially spreading at the same rate.

Lab assays of SARS-CoV-2 have dramatically sped up phenotypic assessment of new variants (Cao et al. 2023). Within days of new variants emerging, information has been shared by groups around the world, evaluating immune evasiveness (e.g., the titer of neutralizing antibodies in convalescent plasma required to prevent infection of cell lines) and efficiency of binding to ACE-2 receptors (e.g., via twitter, @yunlong_cao). These assays often find that infection with one variant (e.g., BA.1) builds higher neutralizing capacity against that variant than other variants (e.g., BA.2), indicating some loss of immune evasiveness following infection with a variant (Cao et al. 2023). The impact on long-term immune evasion and reinfection rates for different variants remains an open question, and one whose answer determines the impact on endemic incidence of disease (Figure 5A,B).

We can counter variant-induced rises in cases, however, by encouraging higher uptake rates of vaccines and other non-pharmaceutical interventions. These measures always help individuals reduce their own risk of infection and the risk of infecting those around them (Figure 7). Widespread, but not universal, uptake is needed to substantially reduce levels of disease (Figure 8), except if the disease is near eradication (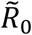 near 1). The benefits could be enhanced by encouraging NPI measures around those who are most at risk of adverse outcomes and in places and times where risks of infection and/or the health care burden are high. Particularly valuable are investments in measures that protect all, regardless of uptake (such as improved air filtration and ventilation, adequate testing and job security to stay home when sick).

The models explored herein lack many important epidemiological details, including spatial and age structure in contact rates and seasonal variation in transmission risk. As such, the results are meant to guide expectations rather than provide precise predictions. Details were sacrificed in an effort to help us better understand how the endemic level of disease is likely to change in the future, in response to our efforts as well as further evolution of the virus.

## Data Availability

All data produced in the present work are contained in the manuscript, as well as accompanying supplementary material (available online)

http://www.zoology.ubc.ca/~otto/Research/Endemic

## Supplementary Materials (FOR REVIEW)

All proofs and code needed to generate the figures are available in *Mathematica* and PDF versions at https://www.zoology.ubc.ca/~otto/Research/Endemic (to be deposited in Dryad).

## Acknowledgements

We thank all the authors, developers, and contributors to the VirusSeq database for making their SARS-CoV-2 sequences publicly available. We thank especially the Canadian Public Health Laboratory Network, academic sequencing partners, diagnostic hospital labs, and other sequencing partners for the provision of the Canadian sequence data used in Figure 1. The authors would like to thank the scientific community of the Coronavirus Variants Rapid Response Network (CoVaRR-Net), particularly Fiona Brinkman, Carmen Lia Murall, Jesse Shapiro, and Justin Yue for helpful discussions about variants. Funding was provided by the Natural Sciences and Engineering Research Council of Canada to SPO (RGPIN-2022-03726), to AM (RGPIN-2022-03113), and to CC (CANMOD; RGPIN-2019-06624), by the Canada Research Chairs Program (SPO and AM) and the Canada 150 Research Chair Program (CC), and by the Canadian Institutes for Health Research (CIHR) operating grant to CoVaRR-Net.

## Appendix 1 Modelling the spread of variants

### Dynamics

We include infections by a resident variant (*I*) and a new variant (*I*^∗^) in the SIR_n_ epidemiological model illustrated in Figure 1. By allowing for multiple recovered classes, we can model new variants that are more immune evasive by allowing them to infect earlier in the waning period (infecting individuals in the last *m* recovered compartments *R*_*j*_), when antibody levels are high enough to prevent infection by the resident virus but not the new variant. The dynamics are then described by the following set of differential equations:

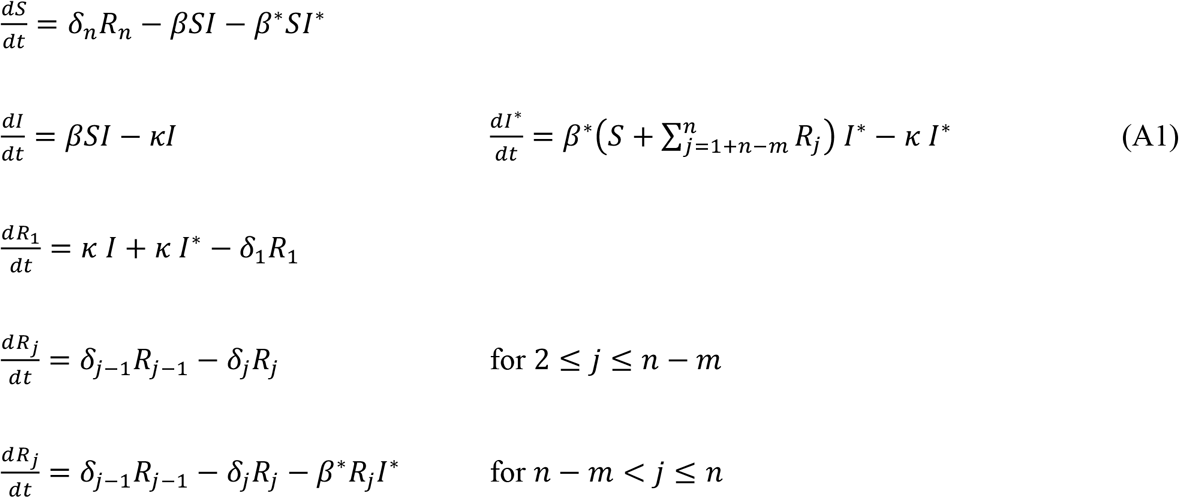

Setting all waning rates between recovered classes equal to *δ*_*i*_ = *δ*/*n* ensures that the average time from first recovering to returning to the susceptible class has a mean of 1/ *δ*days. The distribution of waning times is then given by a gamma distribution with a coefficient of variation (CV) of 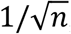, becoming more bell shaped with higher *n* (Hethcote, Stech, and Van Den Driessche 1981).

### Spread of a new variant

The spread of a new variant into a population at the stable endemic equilibrium (equation (2)) is given by the leading eigenvalue, *λ*_*L*_ of the external stability matrix describing the dynamics of the variant (see details in the Supplementary *Mathematica* package), which equals:

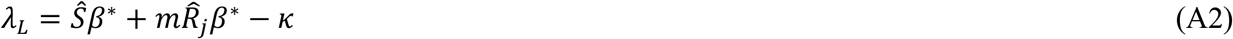

If the new variant did not change the transmission rate (*β*^∗^ = *β*) and was unable to infect any additional sector of the population (*m* = 0), it would be neutral (*λ*_*L*_ = 0, plugging in (2)).

The selection coefficient favouring a new variant is defined by the rise in frequency of the new variant relative to the old variant (^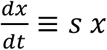^, where *x* =freq(new variant)/freq(old variant)), which predicts an exponential rise in the relative frequency of the new variant over time (*x*_*t*_ = *e*^*s t*^*x*_0_). The strength of selection can thus be estimated empirically by the slope on a logit plot (plotting log of *x*_*t*_ over time). Near the endemic equilibrium, it can be shown that selection, defined in this way, equals *λ*_*L*_ (see Supplementary *Mathematica* package). Plugging in equation (2) for *Ŝ* into (A2) then gives the selection coefficient reported in equation (4).

### Parameter values

We consider the following parameter values for the current endemic phase during which Omicron predominates, giving the nominal value considered typical and the plausible range in square brackets:

- *κ* of 0.2 (mean of 5 days) [range of 3-10 days]. Source: Estimates of the infectious period for Omicron vary depending on the study design, but several studies are consistent with infectiousness for a couple of days prior to symptom onset and five days thereafter (UKHSA 2023). We take into account some self-isolation upon infection and use a five-day average infectious period as a default.
- *δ* of 0.008 (mean of 125 days) [range of 100-180 days]. Source: Waning rate depends on the exact sequence of vaccinations and infections. The half-life of protection against symptomatic infection with Omicron among studies summarized by Menegale et al. (Menegale et al. 2023) was 87 days without a booster and 111 days with a booster, yielding *δ* values ranging from 0.0071 to 0.0094 per day. Waning rates were similar for older and younger individuals (Menegale et al.’s eFigure 14).
- *Î* of 2% [range of 0.5%-4%]. Sources: The last report of the Coronavirus (COVID-19) Infection Survey UK (Office for National Statistics 2023), which assayed nose and throat swabs from households, found 2.66% of England were infected (13 March 2023). In Canada, models suggest that 1 in 28 were infected the week of 16 April 2023, while 1 in 80 were infected the week of 9 September 2023 (COVID-19 Resources Canada 2023).
- Age structure for Canada: 13% of the population is 70+ in age (Statistics Canada 2023).

Combining these estimates with equations (2) and (3) allows estimation of the transmission rate and reproductive number. The nominal parameters given above yield estimates of β = 0.42 and 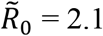, ranging from β = {0.11-2.27} and 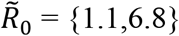 (Table S1), although some combinations are not possible (e.g., a mean waning time of 180 days is inconsistent with an incidence of *Î*= 4% if mean clearance times are too short, *κ* > 0.13), and the disease is then expected to decline.

The expected number of disease bouts per year is 1.46 for the nominal parameter values, ranging from 0 (when the disease disappears) to 2.92 when incidence is high (*Î* = 4%), waning is fast (*δ*= 1/100), and recovery is fast but not so fast that the disease disappears (*κ* = 1/5).

We can also calculate the expected number of infections per year for an individual who is vaccinated at regular intervals (every *T* days). For simplicity, we make the approximation that vaccinations are frequent enough and waning slow enough that we need only consider the chance of one infection between vaccinations. If waning times were exponentially distributed, then the probability of becoming infected in the period between vaccinations would be:

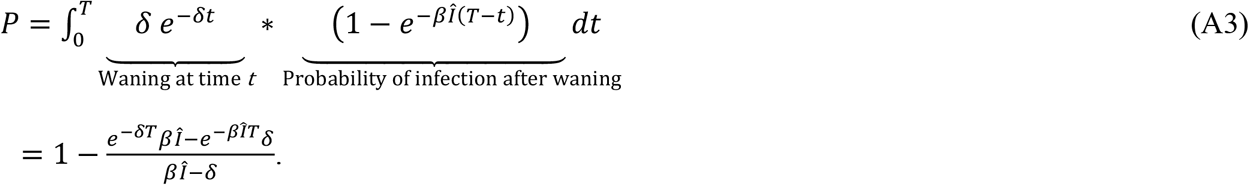

The approximate annual number of infections is then 365 *P*/*T*, which is 0.88 for those on a six-month vaccination interval (*T* = 365/2) and the nominal parameter values (*κ* = 0.2, *δ*= 0.008, *Î*= 2%, *β* = 0.42).

## Appendix 2 Model sensitivity and oscillatory behaviour

Different choices about the number of recovered classes, movement among them, and whether immunity is leaky, as well as the inclusion of a latent period and incomplete seroconversion, were explored to determine sensitivity of the results to model assumptions (Supplementary *Mathematica* file). To simplify the presentation, we focus on the case where daily vaccination rates are low and are ignored (except where noted).

### Alternate models of recovery

In the main text, we used multiple recovered classes in the SIR_n_ model to capture observed declines in neutralizing antibodies, measured on a log scale, over time since vaccination and/or infection. While this reflects the dynamics of neutralizing antibody levels, a side consequence is that the distribution for the total waning time becomes increasingly bell shaped as *n* rises (CV of 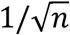). This synchronizes the recovery of individuals infected at the same time. If *n* is large enough, this synchronisation can destabilize the endemic equilibrium, leading to persistent cycles (Hethcote, Stech, and Van Den Driessche 1981). While the rise in frequency of a variant, as described by its selective advantage (equation (4)) and the long-term impact of the variant on the endemic equilibrium (equation (5)) are insensitive to the number of recovered compartments (*n*), the extent of oscillations following the initial spread of the variant are much stronger as *n* increases (Figure S2, panels A-C). Empirically, the distribution of waning times is close to exponential (CV = 1; (Menegale et al. 2023), suggesting that intrinsic oscillations are likely to be damped (like Figure S2A, where CV = 1).

Similar behaviour to Figure S2A is seen in a model with only two recovered classes (*n* = 2), corresponding to high (R_1_) and low (R_2_) antibody levels, where an immune evasive variant (but not the resident virus) can infect the second class. By setting the waning rates to *δ*_1_ = *δ/x* (from R_1_ to R_2_) and *δ*_H_ = *δ*/(1 −*x*) (from R_2_ to S), the equilibrium fraction of recovered individuals in the second class (*x*) can be adjusted to allow for more immune evasive variants, while keeping the average time from first recovering to susceptibility at 1/ *δ* days for the resident virus. This model has a nearly exponential waning time, with rapidly dampening oscillations, for more transmissible variants (Figure S2D), more immune evasive variants (Figure S2E), or both (Figure S2F). The selection coefficient and equilibrium remain unchanged, all else being equal (given by equations (4) and (5), respectively).

### Leaky immunity

In the main text, we considered immunity to be polarized: individuals are either susceptible to infection (*S* compartment) or not (*R*_*j*_ compartments, with *j* depending on the variant). There is evidence, however, that SARS-CoV-2 immunity is leaky, such that high viral exposure can lead to infection for those who would otherwise be immune (Lind et al. 2023). Furthermore, variants may differ in the extent of leaky immune (e.g., (Lind et al. 2023) found higher hazard ratios following close exposure for Delta than for Omicron).

We thus explored variants that increased leakiness of immunity, *ξ*, in the SIR (*n* = 1) and SIR_n_ (*n* = 5) models (exploring the latter numerically only in the Supplementary *Mathematica* file). Incorporating leaky immunity in the SIR model changes the dynamics to:

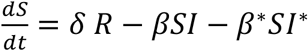

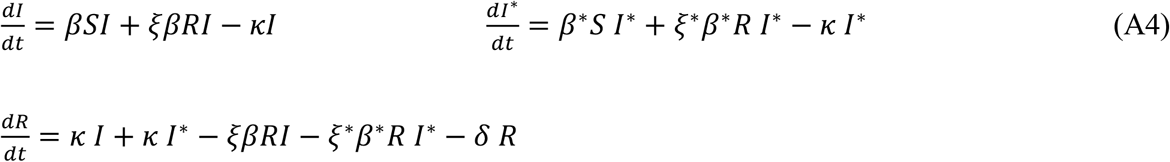

The equilibrium is then:

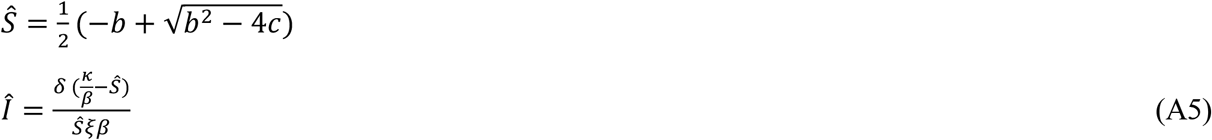

Where 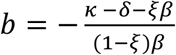 and 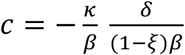. Selection on a variant then becomes:

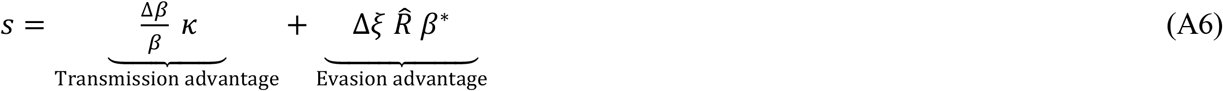

Figure S3 illustrates cases where immunity was robust against the resident virus (*ξ* = 0) but leaky for the variant (*ξ*^∗^ > 0), combined with some to no transmission advantage (panels A to C). Again we see that the same selective advantage (*s*) is consistent with substantially different long-term consequences for endemic disease levels. Variants that exhibit leakier immunity greatly increase the endemic equilibrium, more than seen in Figures 3 and 4 for a given selection coefficient, because all individuals are more prone to infection in the long term, not just those with low antibody levels.

### Latent period

Viral infections are characterized by a latent period between infection and detectible viral load, which is thought to indicate the onset of the infectious period (UKHSA 2023). We can include this period by adding to the SIR_n_ model a latent class (*E*), into which all new infections enter and then exit at rate *ϵ*. Including this period, the equilibrium fraction of infected individuals changes to:

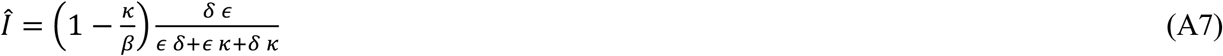

Given that the rate of leaving the latent class is much faster than waning (*ϵ* ≫ *δ*), the last *δκ* term in the denominator is negligible, and *ϵ* cancels out of (A7). Thus, the equilibrium number of infections (*Î*) is nearly unaffected by including a latent class.

Recalculating the leading eigenvalue at this endemic equilibrium, the selection coefficient favoring the new variant (*s* = *λ*_*L*_) changes slightly when a latent period is added, from *s* given by equation (4) to:

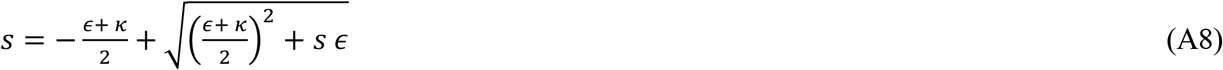

Assuming that the spread of the variant is slow relative to the latent and infectious periods (*s* ≪ *ϵ, κ*), adding a latent period causes selection to weaken slightly, with (A8) approaching 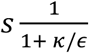. This occurs because only during a fraction 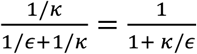 of the generation time of the virus is it infectious. Xin at al. (Xin et al. 2023) estimate a mean latent period of 3.1 days for Omicron. For the parameters considered typical of Omicron (Appendix 1), selection would be ∼70% as strong with a latent period. We ignore this correction to simplify the model presentation.

### Seroconversion

Another real-world complication is that not all individuals seroconvert following infection or vaccination (i.e., not all infections elicit a robust immune response). If a fraction *q* of infections boost immunity (meaning here that they recover to the *R*_1_ compartment in the SIR_n_ model), while 1–*q* return become susceptible again (returning to the *S* compartment), the equilibrium fraction of infected individuals changes to:

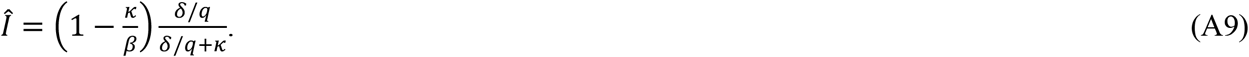

Thus, decreasing the seroconversion rate by a factor *q* has the same effect on the endemic equilibrium as increasing the waning rate by a factor 1/*q* (equation (5)), and the same holds for the selection coefficient of a variant, *s* (equation (4), supplementary *Mathematica* file). The temporal dynamics of the wavelets are slightly different, with immediate waning for those who do not seroconvert and slower waning for those who do (supplementary *Mathematica* file).

Seroconversion rates for vaccines can also be included, changing the equilibrium to:

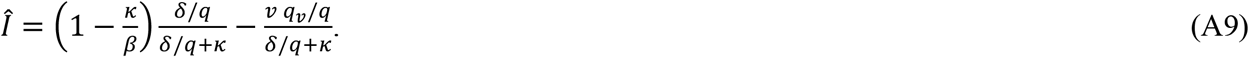

where *q*_*v*_ is the seroconversion rate for vaccination (here meaning the probability that a vaccine dose boosts antibodies and provides protection from infection). If seroconversion rates are similar following infection and vaccination (*q*. = *q*), then the results for selection (equation (3)) and endemic incidence (equation (4)) are again the same if we replace *δ*(ignoring seroconversion) with *δ*/*q* (including it).

To simplify the presentation, we do not explicitly include seroconversion but consider a range of waning rates to cover both seroconversion and waning.

Empirically, high seroconversion rates have been reported following vaccination with a single dose of Pfizer’s BNT162b2 (*q* = 99.5%) or AstraZeneca ChAdOx1 (*q* = 97.1%), leading to antibodies recognizing the spike protein (Wei et al. 2021). Slightly lower seroconversion (*q* = 93.5-95.3%) was observed following infection in early 2020 (Oved et al. 2020). An estimate following Omicron infection inferred even lower rates of seroconversion of *q* = 74-81% (here examining antibodies to nucleocapsid, as anti-spike antibodies were nearly universal in the highly vaccinated population examined; (Erikstrup et al. 2022)).

## Appendix 3 Non-pharmaceutical interventions

We consider an expansion of the SIR_n_ epidemiological model to allow heterogeneity in behaviour. As illustrated in Figure S4, we now allow two classes of individuals, those who regularly adhere to stronger NPI measures, such as masking (indicated by an ‡), and those who do not:

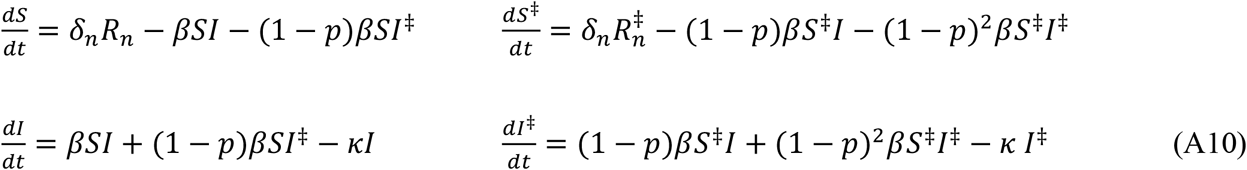

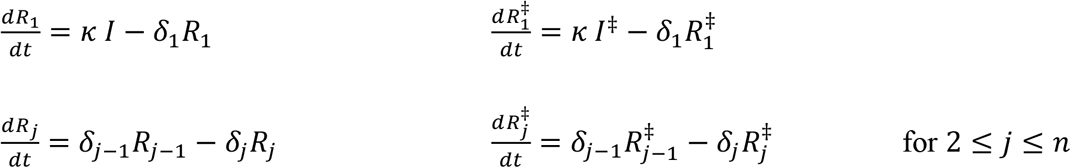

where the last line of equations is repeated for the remaining waning classes (*j* from 2 to *n*). We again set all rates between waning classes to *δ*_D_ = *δ*/*n* (mean waning time of 1/ *δ*days). The new parameter *p* measures the protection provided when one individual in an interaction engages in NPI measures (reducing *β* by a factor 1–*p*). If both infected and susceptible individuals uphold these measures, transmission is reduced by (1– *p*)^2^. All variables are measured as proportions of the total population, with *f* being the fraction of the population carrying out NPI measures, such as masking (the sum of the ‡ variables).

There are two equilibria of this system (A10), one where the disease is absent and one where the disease is endemic at:

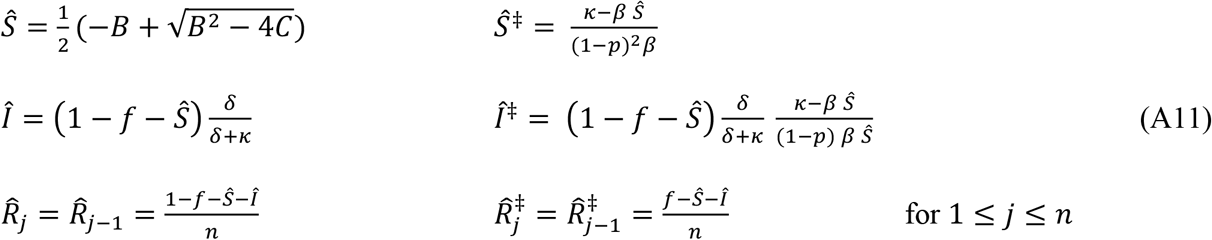

where 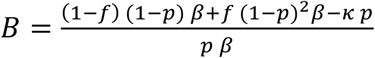 and 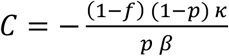. The disease-absent equilibrium is locally stable when transmission rates are low relative to clearance, such that the endemic reproductive number if everyone were susceptible is less than one, 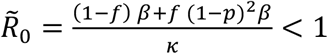 1, in which case the endemic equilibrium does not exist (i.e., not all variables are positive). Otherwise, when 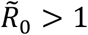, the endemic equilibrium exists and is stable for the parameters considered, but it may become unstable for large *n* (Hethcote, Stech, and Van Den Driessche 1981).

At this equilibrium, the risk that an individual is in the infected class at any point in time is *Î*^‡^/*f* if they regularly mask and *Î*/(1 −*f*) if they do not, from which we calculate the relative risk in the main text. The population-level impact of NPI measures, such as masking, is determined by analysing the fraction of the population expected to be infected at any point in time, *Î* + *Î*^‡^.

The above assumes that an individual’s choice about engaging in NPI measures remains constant over time, but we also consider the opposite case (detailed in the Supplementary *Mathematica* file), where individuals rapidly switch between engaging or not in NPI measures. Assuming that the behaviour persists over the short time frame of an infection but that individuals switch often while in the longer susceptible or recovered phases, we can simplify the model by monitoring only those engaging in NPI measures at the time of exposure, with *f* then representing the probability that an individual engages in the NPI measures at that time. We thus only sub-divide the infectious class into those who were or were not practicing the NPI measures at the time of infection (*I*^‡^ or *I*, respectively). The dynamics are then:

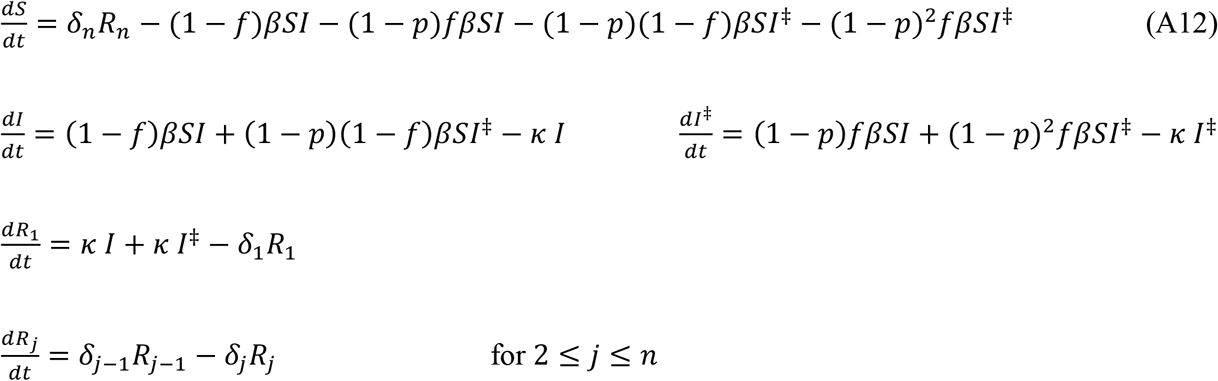

Results using (A12) instead of (A10) are similar, except that practicing and non-practicing individuals are equally likely to be susceptible at the time of exposure, so that the individual-level protective effect of the NPI measure now depends only on *p* and not on 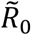, as discussed in the text.

### Parameters

The protection (*p*) and uptake (*f*) depend on the NPI measure considered (The Royal Society 2023). Here we briefly review data on masking as a protective measure. One metaanalysis of randomized control studies prior to the COVID-19 pandemic found protection provided by masks was *p* = 16% for respiratory infections, rising to *p* = 24% in studies longer than two weeks (Li et al. 2022). Importantly, many individual studies were underpowered but the results were consistent across studies (see Figure 2 in Li et al. 2022).

For COVID-19, a metaanalysis of the impact of mask mandates estimated a 25% reduction in transmission rates, comparing transmission levels predicted if everyone were in the class that self-report wearing masks “most of the time in some public places” to that if no one wore masks (Leech et al. 2022). Importantly, the authors showed that the lifting or imposition of mandates rarely had dramatic immediate effects on mask wearing, emphasizing that mandates are a poor proxy for mask wearing. Their analysis thus benefited from a global analysis of trends in mask-wearing behavior around the time of mandates by incorporating data from a survey of masking behaviour among nearly 20 million individuals.

As argued by Leech et al. (2022), this effect size is likely to be underestimated for a number of reasons. First, the study period (1 May – 1 September 2020) occurred when cloth masks predominated, because high-quality masks were largely unavailable outside of health care settings. Second, the definition of mask use was broad and included individuals who only occasionally mask and do so in few public places. We thus consider that *p* = 0.25 represents a lower bound on the protection provided by masking.

Higher values are plausible when using high-quality masks and doing so consistently in indoor public spaces. For example, masks provided a stronger benefit, reducing the odds ratio of infection by an average of 50% among the studies summarized within healthcare settings (The Royal Society 2023). We thus consider *p* = 0.5 to represent a reasonable upper bound on the protection provided by masking attainable by consistent wearing of high-quality masks. Combinations of NPI measures, including improved ventilation, avoiding crowded indoor environments, testing and self-isolation, and masking may provide considerably stronger protection.

**Table S1:** Parameter estimates and inferred rates. The first three columns are estimated from the literature (Appendix 1). Using equation (2), these parameters provide estimates for the transmission rate (*β*), the endemic reproductive number 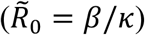, the fraction of susceptible individuals (*Ŝ*), and the estimated annual number of infections at the endemic equilibrium (inferred values are in italics). The first row gives the nominal parameter values used in the text. “NA” are parameter combinations that do not sustain an endemic equilibrium of the disease. Estimates are given without ongoing vaccination (*v* = 0) but are nearly identical with low rates, as in Canada during the summer of 2023 (*v* = 0.00012, all differences <12%; see supplementary *Mathematica* file for estimates with higher rates of vaccination).

| Incidence<br>($\hat{I}$) | Waning rate<br>($\delta$) | Recovery<br>rate ($\kappa$) | Transmission<br>rate ($\beta$) | Reproductive<br>number ($\tilde{R}_0$) | Susceptible<br>($\hat{S}$) | Annual # of<br>infections |
| --- | --- | --- | --- | --- | --- | --- |
| **0.02** | **0.008** | **0.2** | ***0.417*** | ***2.083*** | ***0.48*** | ***1.46*** |
| 0.02 | 0.008 | 0.333 | *2.273* | *6.818* | *0.147* | *2.433* |
| 0.02 | 0.008 | 0.1 | *0.137* | *1.37* | *0.73* | *0.73* |
| 0.02 | 0.01 | 0.2 | *0.345* | *1.724* | *0.58* | *1.46* |
| 0.02 | 0.01 | 0.333 | *1.064* | *3.191* | *0.313* | *2.433* |
| 0.02 | 0.01 | 0.1 | *0.128* | *1.282* | *0.78* | *0.73* |
| 0.02 | 0.006 | 0.2 | *0.769* | *3.846* | *0.26* | *1.46* |
| 0.02 | 0.006 | 0.333 | NA | NA | NA | NA |
| 0.02 | 0.006 | 0.1 | *0.161* | *1.613* | *0.62* | *0.73* |
| 0.005 | 0.008 | 0.2 | *0.23* | *1.149* | *0.87* | *0.365* |
| 0.005 | 0.008 | 0.333 | *0.424* | *1.271* | *0.787* | *0.608* |
| 0.005 | 0.008 | 0.1 | *0.107* | *1.072* | *0.932* | *0.182* |
| 0.005 | 0.01 | 0.2 | *0.223* | *1.117* | *0.895* | *0.365* |
| 0.005 | 0.01 | 0.333 | *0.402* | *1.207* | *0.828* | *0.608* |
| 0.005 | 0.01 | 0.1 | *0.106* | *1.058* | *0.945* | *0.182* |
| 0.005 | 0.006 | 0.2 | *0.245* | *1.227* | *0.815* | *0.365* |
| 0.005 | 0.006 | 0.333 | *0.48* | *1.439* | *0.695* | *0.608* |
| 0.005 | 0.006 | 0.1 | *0.11* | *1.105* | *0.905* | *0.182* |
| 0.04 | 0.008 | 0.2 | NA | NA | NA | NA |
| 0.04 | 0.008 | 0.333 | NA | NA | NA | NA |
| 0.04 | 0.008 | 0.1 | *0.217* | *2.174* | *0.46* | *1.46* |
| 0.04 | 0.01 | 0.2 | *1.25* | *6.25* | *0.16* | *2.92* |
| 0.04 | 0.01 | 0.333 | NA | NA | NA | NA |
| 0.04 | 0.01 | 0.1 | *0.179* | *1.786* | *0.56* | *1.46* |
| 0.04 | 0.006 | 0.2 | NA | NA | NA | NA |
| 0.04 | 0.006 | 0.333 | NA | NA | NA | NA |
| 0.04 | 0.006 | 0.1 | *0.417* | *4.167* | *0.24* | *1.46* |

**FIGURE S1:**
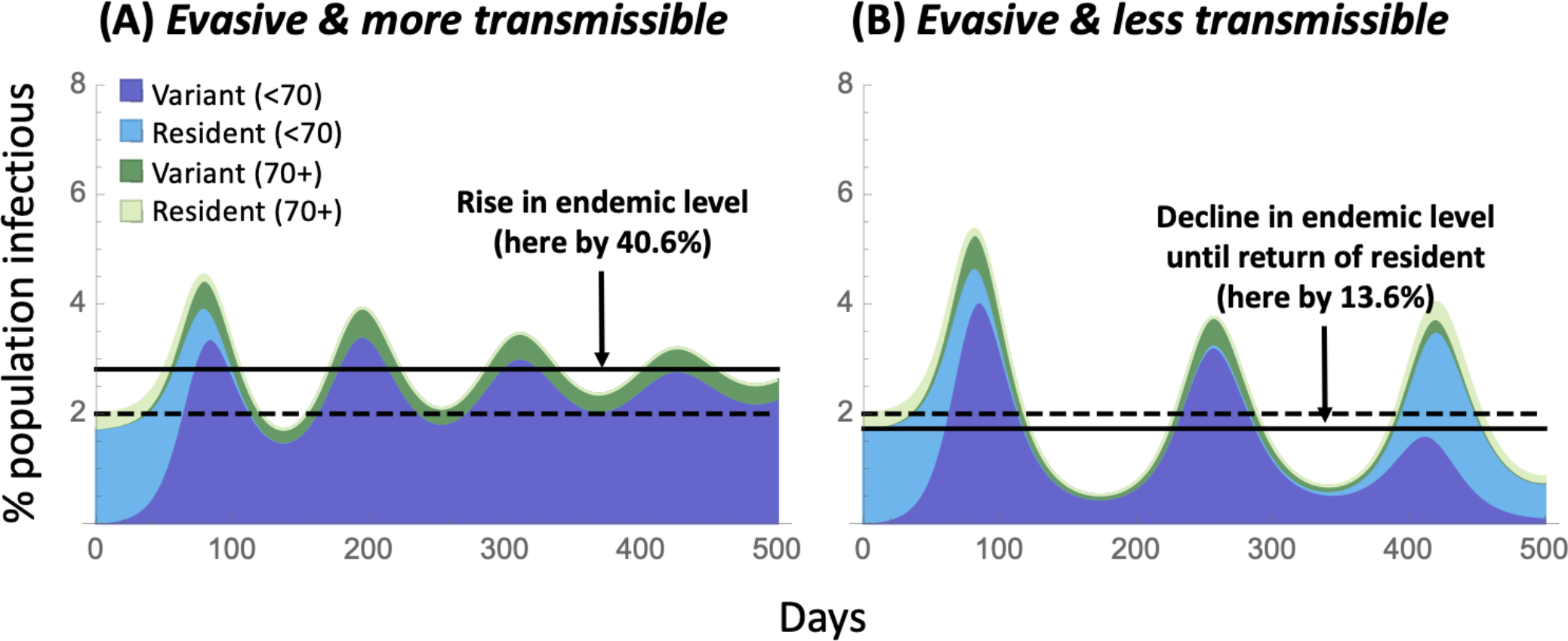
Variants combining immune evasiveness and changes in transmissibility. Panel A: Variant is persistently immune evasive (by half the amount shown in Figure 3B, able to infect *m* = 1 out of *n* = 5 recovered classes) and more transmissible (increasing *β*, and hence 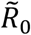, by 17%). Panel B: Variant is even more immune evasive (50% more than in Figure 3B, able to infect *m* = 3 out of *n* = 5 recovered classes) and less transmissible (decreasing *β*, and hence 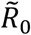, by 17%), with immune evasion being transient. In both cases, *β*^∗^ was chosen to give the variant the same selective advantage as in Figure 3 (*s* = 8.3%), leading to the same initial rate of spread of the variant (dark shading) in a resident population of viruses (light shading). Because the variant is only transiently immune evasive in Panel B, the resident lineage, which is more transmissible, eventually takes over. Parameters: *κ* = 0.2, *δ*= 0.008, *Î* = 2%, *β* = 0.42. the nominal parameter estimates given in Appendix 1 for all age classes.

**FIGURE S2:**
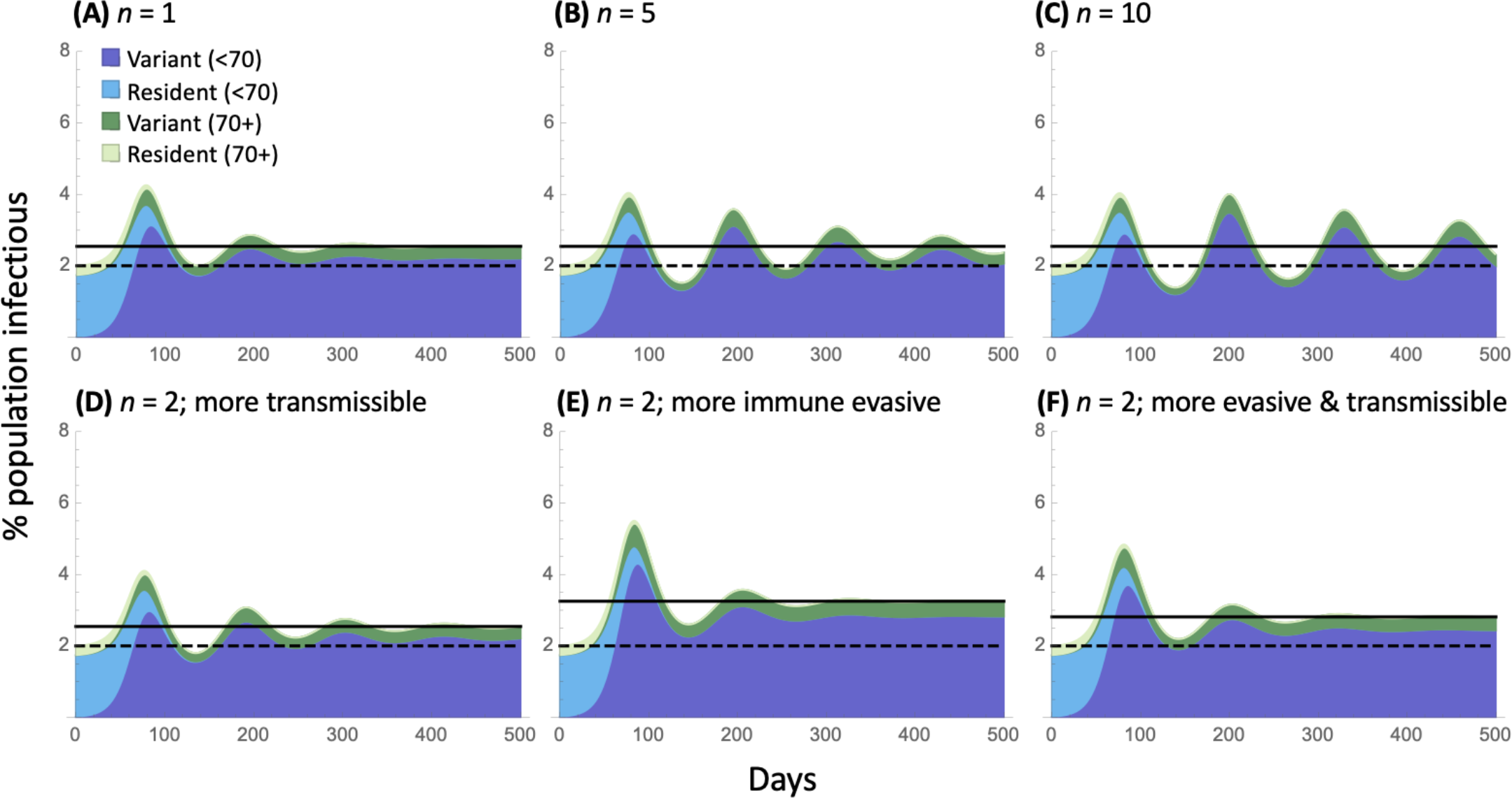
Robustness to model structure. Panels (A)-(C) illustrate the sensitivity of the dynamics to the number of recovery classes (*n*) for a more transmissible variant that increases *β* by 42% (panel B is equivalent to Figure 3). Holding the expected waning period constant at 1/*δ*= 125 days, increasing the number of compartments causes a more bell-shaped distribution of the waning period and increases oscillations following the spread of the variant (from panel A to C). Panels (D)-(F) consider two recovered compartments (*n* = 2), corresponding to high and low neutralizing antibody levels. At the endemic equilibrium, either 40% (*x* = 0.4; panels D, E) or 20% (*x* = 0.2, panel F) of recovered individuals are in the low-immunity compartment, which can be infected by a more immune evasive variant (panels E and F). The variant in panel (D) is only more transmissible. In each case, the transmissibility of the variant is adjusted to hold its selective advantage constant at (*s* = 8.3%), increasing *β* by (A)-(D) 42%, (E) 0%, and (F) 17%. Parameters: *κ* = 0.2, *δ*= 0.008, *Î*= 2%, *β* = 0.42, the nominal parameter estimates given in Appendix 1 for all age classes.

**FIGURE S3:**
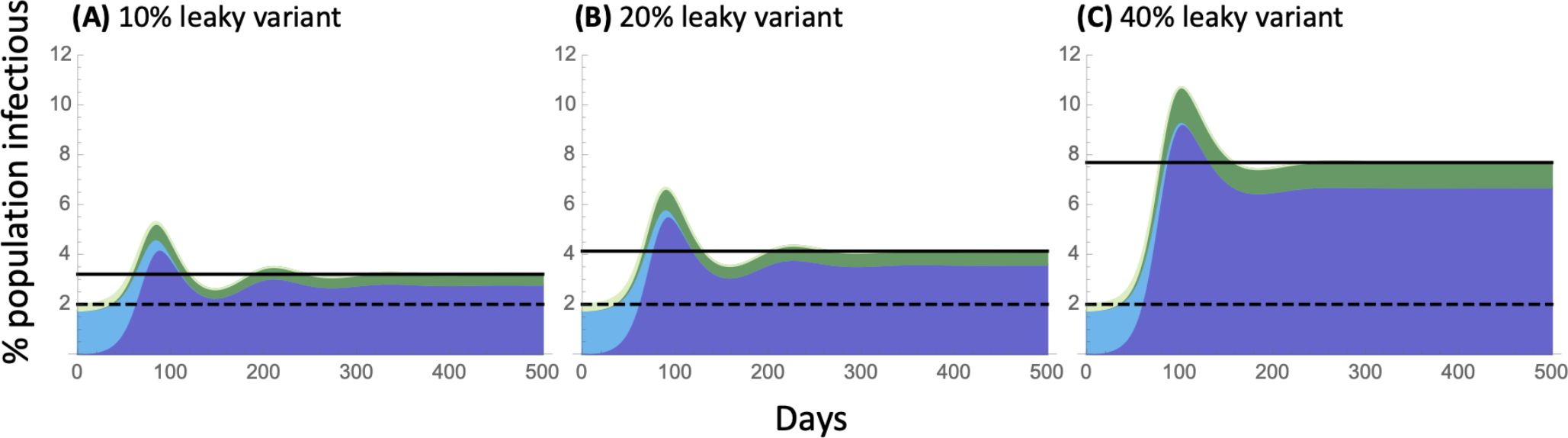
Robustness to model structure: allowing leaky immunity. Panels illustrate the standard SIR model (*n* = 1) when variants increase leakiness of immunity. Individuals infectious with the variant can infect all recovered individuals with a transmission rate that is (A) *ξ* = 10%, (B) 20%, and (C) 40% times *β*, whereas individuals carrying the resident virus can only infect susceptible individuals (*ξ* = 0). In each case, the transmissibility of the variant is adjusted to hold its selective advantage constant at *s* = 8.3%, which required increasing *β* by (A) 28%, (B) 17%, and (C) 0%. Note that the y-axis has been increased relative to previous figures due to the greater long-term impact of variants able to infect all recovered classes by increasing the leakiness of immunity. Parameters: *κ* = 0.2, *δ*= 0.008, *Î* = 2%, *β* = 0.42, the nominal parameter estimates given in Appendix 1 for all age classes.

**FIGURE S4:**
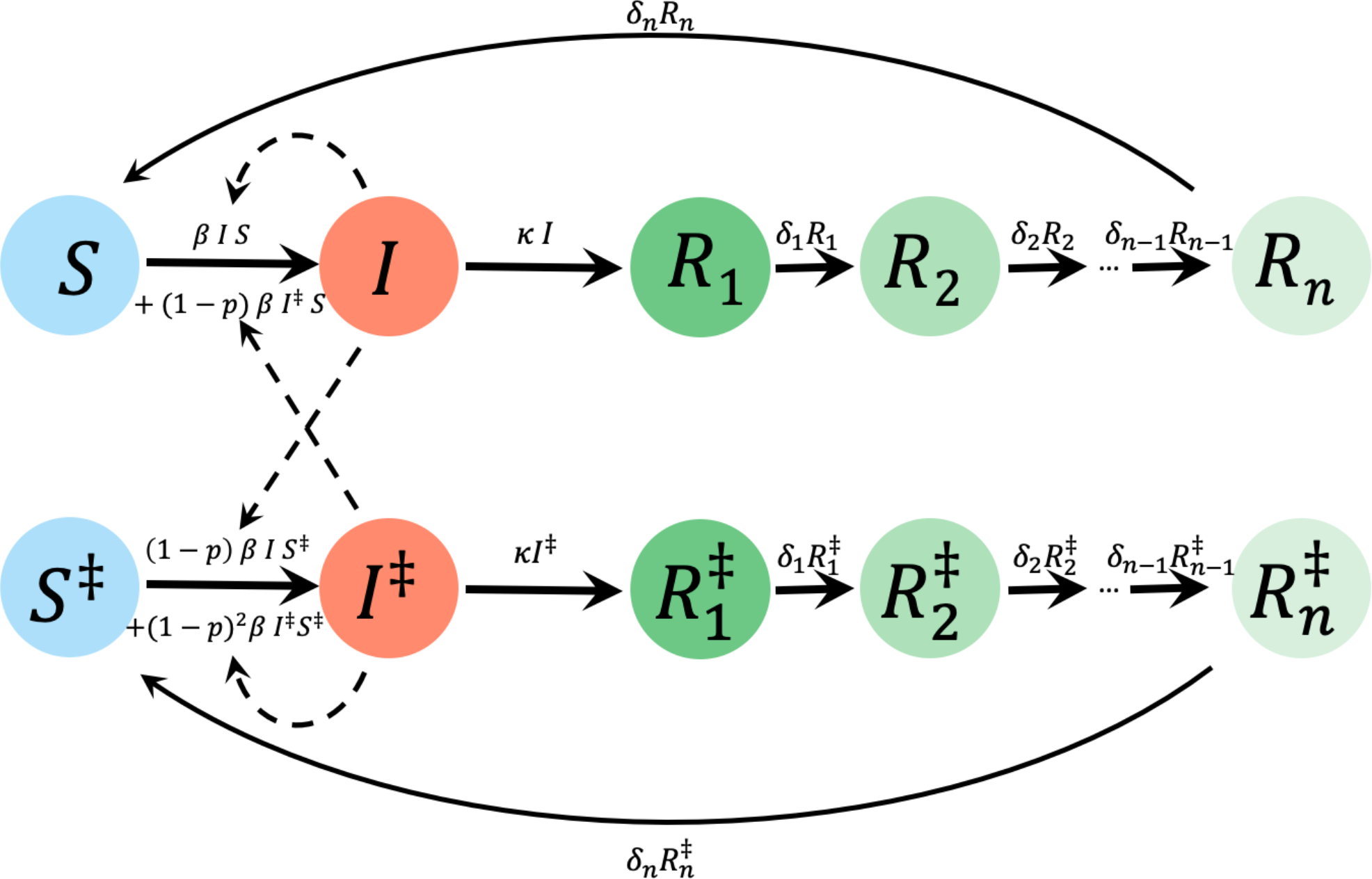
Epidemiological model used to explore the benefits of NPI measures, such as masking. We include behavioural heterogeneity in SIR_n_ model, where variables with a ‡ denote individuals engaging in the NPI measure(s). As before, *S* are susceptible individuals, *I* are infectious, and *R*_*n*_ are recovered, with immunity at different stages of waning. The fraction of individuals engaging in NPI measures is 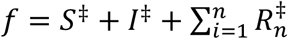. See Supplementary *Mathematica* file for an alternative model where individuals rapidly switch behaviours (switching between the upper and lower row of circles). Parameters are *β*: transmission rate, *p*: protection provided by masking (modeled as a reduction in transmission by a factor (1-*p*) for each person in an interaction wearing a mask), *κ*: recovery rate, and *δ*_*j*_: per-class waning rate.

